# Generating synthetic tau-PET scans in Alzheimer’s disease from MRI, blood biomarkers and demographics with deep learning

**DOI:** 10.64898/2026.05.06.26352540

**Authors:** Linda Karlsson, Olof Strandberg, Ruben Smith, Weizhong Tang, Ida Arvidsson, Kalle Åström, Kevin Oliveira Hauer, Shorena Janelidze, Erik Stomrud, Sebastian Palmqvist, Philip B. Verghese, Joel B. Braunstein, Alzheimer’s Disease Neuroimaging Initiative, PREVENT-AD Research Group, Gregory Klein, Sergey Shcherbinin, William J. Jagust, Sylvia Villeneuve, Renaud La Joie, Gil D. Rabinovici, Niklas Mattsson-Carlgren, Jacob W. Vogel, Oskar Hansson

## Abstract

Tau protein aggregation in the brain is a hallmark of Alzheimer’s disease (AD). Positron emission tomography (PET) is the only in vivo method to visualize tau pathology and estimate both its burden and regional distribution, but the use of tau-PET is constrained by high cost and limited accessibility. Here, we develop a deep learning model to synthesize tau-PET scans from more accessible data: structural magnetic resonance imaging (MRI), demographics, and when available, blood biomarkers. We included 5,191 participants across the AD continuum or with another neurological disorder from 13 cohorts (mean age 70 years, 51% female) and optimized a 3D U-Net neural network with residual and attention units for this task. In held-out test data, synthetic tau-PET reliably modeled tau burden, with correlations of R=0.77–0.86 with true tau-PET across individuals in common AD regions of interest. Spatial similarity between synthetic and true tau-PET was likewise high, with mean regional correlation of R=0.75. Synthetic scans also captured clinically meaningful prognostic information comparable to true tau-PET, including distinction between early (HR=12, p<0.001) and late (HR=45, p<0.001) stages of tau accumulation. These findings demonstrate that clinically informative synthetic tau-PET scans can be generated from widely available modalities using deep learning, potentially offering a scalable and cost-effective approach for estimating tau AD pathology in the brain.

## Introduction

Alzheimer’s disease (AD) is the most common cause of dementia, affecting millions of people worldwide.^1^ In AD, pathological accumulation of proteins occurs in the brain, including amyloid-β (Aβ), which forms extracellular plaques, and tau, which forms intracellular neurofibrillary tangles.^2^ While Aβ accumulation marks the earliest (preclinical) disease stages, the later-occurring tau pathology is more closely linked to neurodegeneration and clinical symptoms, underscoring its importance for disease staging and prognosis.^3^ Methods to detect and quantify AD-related tau pathology include cerebrospinal fluid and blood-based biomarkers, whereas MRI can capture complementary information on downstream neurodegeneration and atrophy patterns.^4,5^ Yet, the only *in vivo* technique that can accurately assess both the load and spatial distribution of insoluble tau aggregates is PET imaging. Tau-PET has been shown to increase diagnostic confidence of physicians^6,7^, to capture distinct patterns of disease heterogeneity related to clinical presentation and prognosis in AD^8–11^, and to help differentiate AD from other neurodegenerative disorders^12–14^. Furthermore, it uniquely enables *in vivo* staging of tau pathology according to regional burden and distribution, for example according to the commonly used Braak staging scheme ^15,16^, allowing direct assessment of disease progression. Despite its clear value, tau-PET remains currently largely unavailable to most patients and memory clinics worldwide due to its high cost and limited radiotracer access. With the emergence of disease-modifying therapies for AD, including recently approved anti-amyloid treatments^17,18^ and tau-targeting drugs in development^19^, the need for reliable, accessible, and scalable diagnostic and staging tools has become increasingly evident. To ensure timely access to treatment while minimizing burden on healthcare systems, strategies are needed that reduce reliance on costly imaging techniques such as PET, without compromising the ability to capture both the load and spatial pattern of pathology.^20^

A promising strategy to reduce reliance on PET while still approximating its unique information is to use artificial intelligence (AI) to generate synthetic tau-PET from other neuroimaging modalities. However, high performance of AI based approaches has typically only been achieved when using other PET modalities as input, and thereby not eliminating the need of a costly PET scan in the diagnostic workflow. For example, Lee et al. successfully synthesized tau-PET scans from fluorodeoxyglucose (FDG)-PET and Aβ-PET, but performance was notably reduced when only using MRI.^21^ Similarly, Chen et al. showed that full-dose PET images could be predicted from MRI and ultra-low-dose tau-PET, reducing radiation exposure but still requiring a PET scan.^22^ These studies highlight both potential and limitations of generating synthetic tau-PET from other neuroimaging modalities, motivating further investigation of this strategy using larger datasets and additional data types.

Another common strategy is to predict tau burden in specific brain regions rather than generating full images. This approach has been tested using various combinations of input features (e.g. fluid biomarkers^23,24^, multi-modal clinical and MRI data^25^, Aβ-PET, MRI and *APOE* genotype^26^), producing good predictive performance, while also providing insight into how different modalities relate to tau-PET. However, such region-wise or composite predictions have limited descriptive detail, as they reduce rich spatial information into single summary values. Although this simplification can facilitate interpretation, a single composite is usually not sufficient to capture both early and late tau pathology^4,15^, and more details on the spatial tau pattern can provide information relevant for more precise diagnosis and prognosis^9,10,27,28^. Predicting multiple tau-PET composites simultaneously partly addresses this issue, but because the signals in these composites often are correlated, it remains difficult to disentangle whether the predictions truly capture region-specific signals or merely reflect a scaled version of a global tau pattern. To address the last issue, we recently examined the feasibility of separately predicting tau-PET load and spatial information as two distinct outputs from more accessible variables using traditional machine learning (ML) models.^29^ Plasma p-tau217 was the strongest predictor of tau load, whereas MRI features best captured spatial information (hemispheric asymmetry in tau deposition). The findings highlight the potential of combining blood biomarkers and MRI to estimate both tau load and spatial patterns simultaneously.

Building on this foundation, the objective of the present study was to move beyond predicting regional composites and instead generate full synthetic tau-PET scans using deep learning models trained on multiple accessible data modalities (blood biomarkers, MRI and clinical variables). Our aim was to study the performance achievable when training on a relatively large dataset, evaluate whether the synthetic images provide clinically meaningful information, and better understand strengths and limitations of synthetic scans across neurodegenerative diseases and cognitive stages. We gathered data from multiple AD cohorts (totaling n=5,191 participants) and systematically compared different deep learning architectures and input feature configurations to find an optimized model. Using left-out test data, we evaluated image quality compared to true PET using both quantitative and qualitative assessments. We also examined whether the synthetic tau-PET scans provided clinically meaningful information across several diagnostic and prognostic contexts, and whether adding the more tau-specific fluid biomarker eMTBR-tau243 could further improve tau-PET synthesis.

## Results

### Study design

We assembled tau-PET, MRI, demographic information, and (when available) plasma biomarkers across 13 cohorts (see Methods for details). To ensure robust and favorable training conditions, in-house preprocessing of all imaging data was performed (see Methods for details). The final multi-cohort dataset consisted of 5,191 unique individuals (mean [std] age 70 [9.9] years, 51% female, Table 1, Supplementary Table 1), of whom all had tau-PET, T1-weighted MRI and demographic data, and 2,352 had plasma p-tau217 measurements. Participants represented a broad spectrum of clinical diagnoses, including cases with normal cognition (NC, 47.4%), subjective cognitive decline (SCD, 7.53%), mild cognitive impairment (MCI, 19.0%), AD dementia (15.2%), non-AD dementia (4.71%) and other neurological disorders (6.18%). Overall, 51.4% of the individuals were Aβ-positive based on CSF or PET. Data from the multi-cohort dataset was randomly split into train (n_tot_=3,815, n_p-tau217_=1,708), validation (n_tot_=698, n_p-tau217_=326) and test (n_tot_=678, n_p-tau217_=318) sets (participants from all 13 cohorts were represented in all sets). The train and validation sets were used to optimize and train deep learning models, and the test set only for evaluation of the final model. See an overview of the study design in Figure 1.

**Figure 1:**
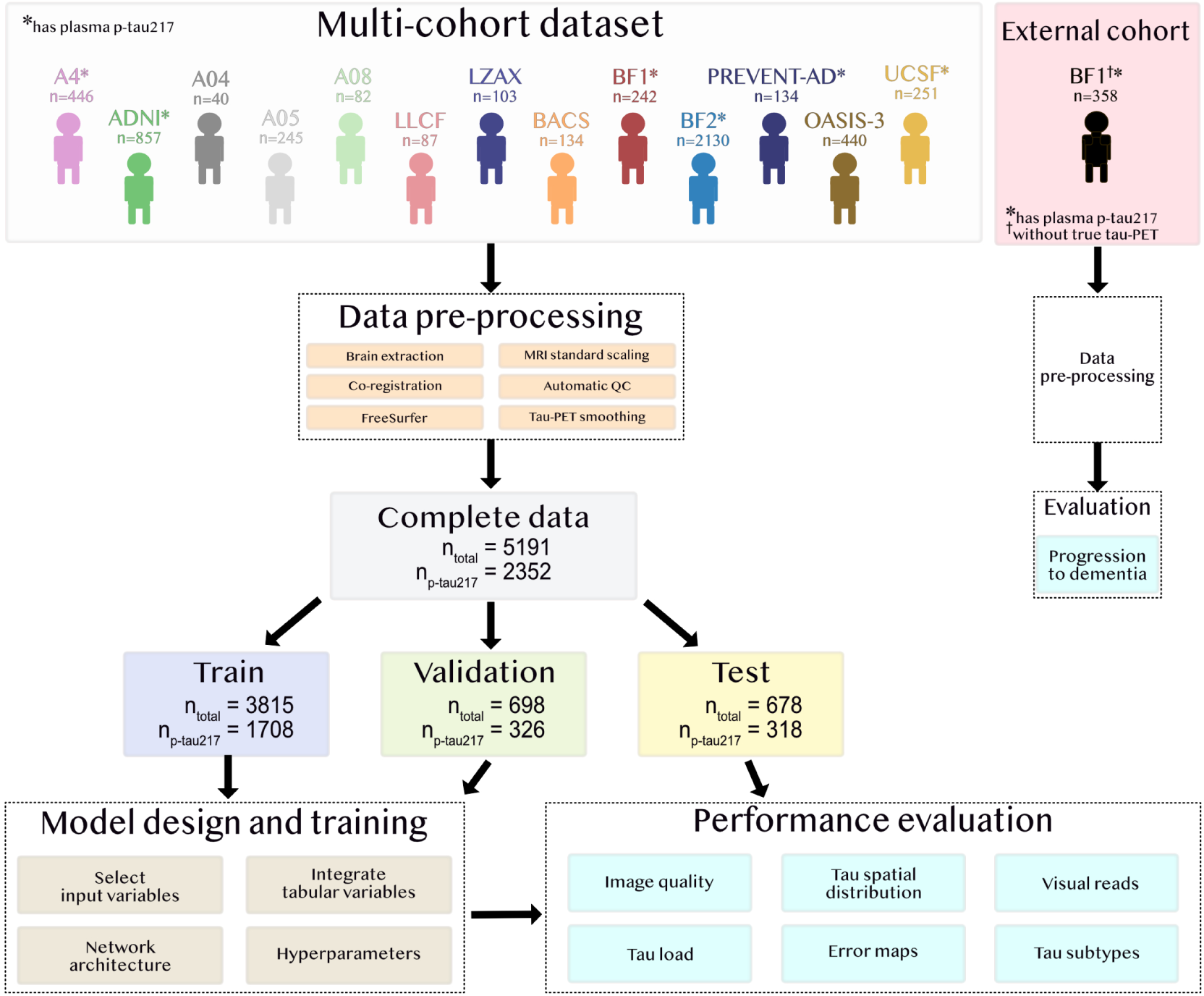
Overview of multi-cohort dataset and study design. The same training, validation, and test split was kept throughout all analyses in this work. The external cohort BF1 consisted of 358 study participants who never underwent true tau-PET, in contrast to the 242 BF1 participants included in the multi-cohort dataset.

**Table 1:** Participant demographics. Characteristics of the multi-cohort dataset. See Supplementary Table 1 for corresponding information stratified by cohort.

|  | **Train set** | **Validation set** | **Test set** | **All** |
| --- | --- | --- | --- | --- |
| **Number of participants** | 3815 | 698 | 678 | 5191 |
| **Number of participants with plasma p-tau217** | 1708 | 326 | 318 | 2352 |
| **Age, mean (std) years** | 70.1 (9.86) | 69.5 (9.74) | 70.2 (10.5) | 70.0 (9.93) |
| **Sex n (%) female** | 1964 (51.5) | 360 (51.6) | 329 (48.5) | 2651 (51.1) |
| **Education level, mean (std) years** | 14.4 (3.77) | 14.6 (3.74) | 14.4 (3.76) | 14.5 (3.67) |
| **APOE ε4 alleles\*, n (%)** |  |  |  |  |
| 0 | 1807 (56.8) | 304 (52.1) | 334 (58.4) | 2445 (56.4) |
| 1 | 1127 (35.5) | 233 (40.0) | 187 (32.7) | 1547 (35.7) |
| 2 | 245 (7.71) | 46 (7.89) | 51 (8.92) | 342 (7.89) |
| **Clinical diagnosis\*, n (%)** |  |  |  |  |
| NC | 1759 (47.5) | 322 (47.5) | 311 (47.0) | 2392 (47.4) |
| SCD | 283 (7.64) | 47 (6.93) | 50 (7.56) | 380 (7.53) |
| MCI | 681 (18.4) | 144 (21.2) | 132 (20.0) | 957 (19.0) |
| AD dementia | 574 (15.5) | 85 (12.5) | 107 (16.2) | 766 (15.2) |
| Non-AD dementia | 176 (4.75) | 37 (5.56) | 25 (3.78) | 238 (4.71) |
| Other neurological disorder | 233 (6.29) | 43 (6.34) | 36 (5.45) | 312 (6.18) |
| **Aβ positive\*, n (%)** | 1811 (51.1) | 334 (51.5) | 341 (52.9) | 2486 (51.4) |
| **Tau-PET tracer, n (%) ([$^{18}$F]floritaucipir / [$^{18}$F]RO948)** | 2270 (59.5) / 1545 (40.5) | 424 (60.7) / 274 (39.3) | 367 (54.1) / 311 (45.9) | 3061 (59.0) / 2130 (41.0) |
| **Plasma p-tau217\*, mean (std) pg/ml** | 0.346 (0.814) | 0.293 (0.216) | 0.319 (0.265) | 0.335 (0.706) |

Non-AD dementia includes Frontotemporal dementia, Lewy body dementia, Vascular dementia, and Posterior cortical atrophy. Other neurological disorder includes Parkinson's disease, Progressive supranuclear palsy, Corticobasal syndrome, multiple system atrophy, traumatic brain injury, chronic traumatic encephalopathy, and bipolar disorder.
Abbreviations: normal cognition (NC), subjective cognitive decline (SCD), mild cognitive impairment (MCI), Alzheimer's disease (AD), amyloid-β (Aβ ).
\*missing data for some participants

### Systematic model optimization

To identify a high-performing deep learning framework, we systematically compared several input variable configurations and convolutional neural network model architectures. Model optimization was done by minimizing the validation loss. We first trained a compact 3D U-Net (∼6M trainable parameters) using only T1-weighted MRI as input. Additional accessible predictors, including blood biomarkers and age and sex variables, were then incorporated one at a time. Less readily available variables like clinical diagnosis and Aβ-status were not included as input. Performance gains were observed when adding age or plasma p-tau217, whereas sex did not contribute to a distinct improvement (Supplementary Table 2). We therefore combined MRI, age, and plasma p-tau217 into a joint model. To ensure flexibility when plasma data were unavailable, missing values were imputed to the training set plasma p-tau217 mean value (a non-informative value), allowing the model to process such cases without excluding participants. We next evaluated different ways of integrating tabular variables, with the best results achieved when age and p-tau217 were added as separate channels at the U-Net bottleneck. Finally, we scaled up the model by increasing network depth and number of filters, and systematically compared different encoder-decoder block architectures. The resulting final model was a 3D U-Net with residual units in the encoder/decoder, an attention unit in the bottleneck, and ∼110M trainable parameters in total (Figure 2). Further details of the optimization procedure are provided in Methods and Supplementary Table 2. The final model and code will be made available at https://github.com/DeMONLab-BioFINDER/karlsson_synthetic_taupet.

**Figure 2:**
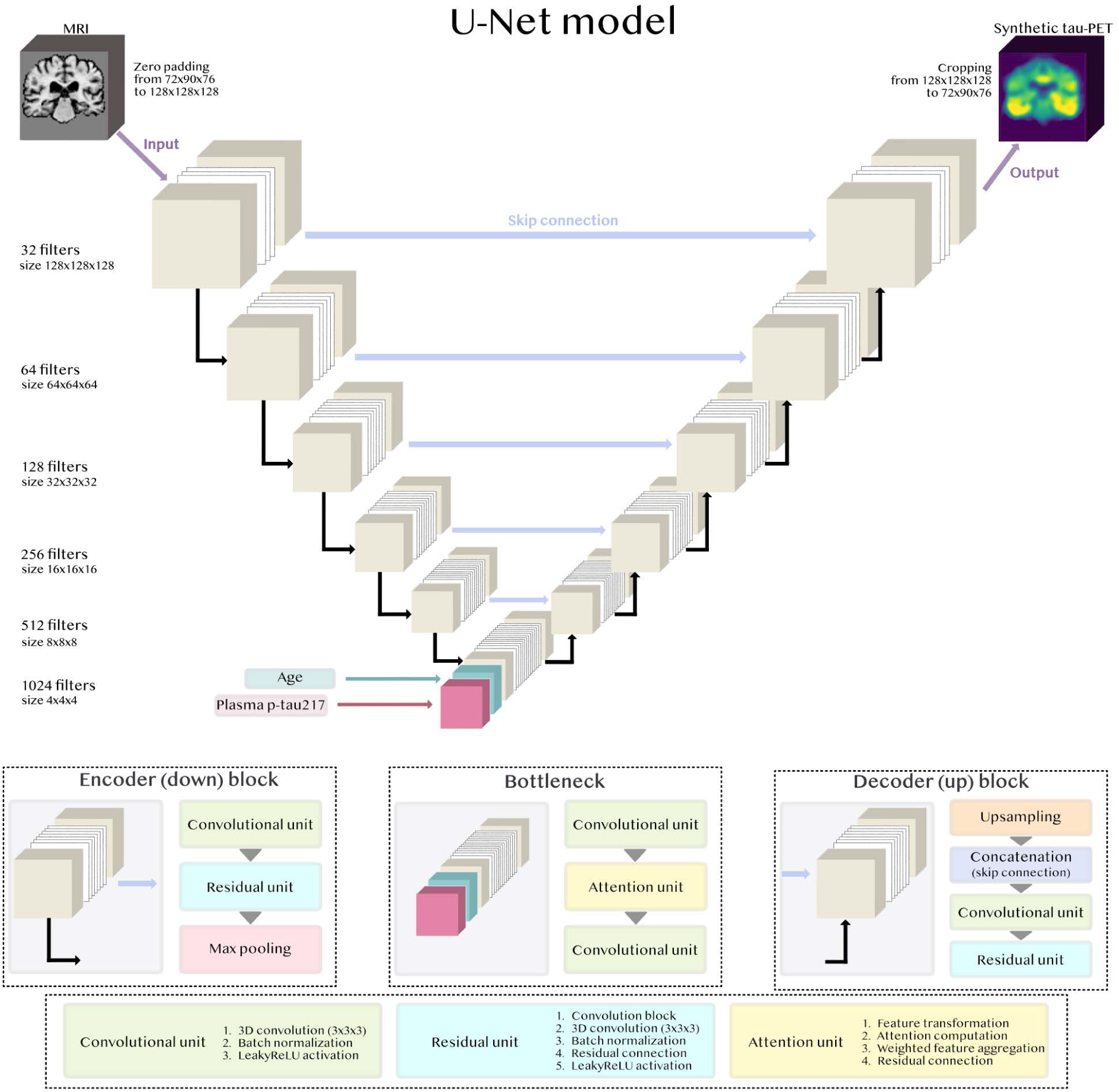
Architecture of final deep learning 3D U-Net model to generate synthetic tau-PET. The final architecture with residual units in the encoder/decoder and an attention unit in the bottleneck. T1-weighted structural MRI was the initial input, and plasma p-tau217 and age were integrated as separate channels in the bottleneck.

### Performance evaluation

Performance was evaluated on tau-PET (standardized uptake value ratio [SUVR] images) from the held-out test set for two different input variable configurations. The first input to the model was MRI, age, and plasma p-tau217, representing the best-case scenario when both imaging and blood biomarkers are available (n=318 in test set). The second excluded plasma p-tau217, leaving only MRI and age as inputs (n=678 in test set), representing a more constrained setting.

#### Synthetic tau-PET scans show adequate image quality

Image quality was evaluated using voxel-wise mean absolute error (MAE) and structural similarity index measure (SSIM) between true and synthetic tau-PET. Average error was low (MAE_avg_=0.075 SUVR for MRI, age & plasma; MAE_avg_=0.074 SUVR for MRI & age) but increased with higher tau burden (Supplementary Figure 1, Supplementary Table 3). This likely reflects the high proportion of cognitively unimpaired Aβ-negative individuals in the multi-cohort dataset, and the lower difficulty of reconstructing low-signal scans (simply producing a uniform, low-intensity signal can still yield relatively low MAE). In contrast, structural similarity (SSIM_avg_=0.81 for MRI, age & plasma; SSIM_avg_=0.81 for MRI & age, Supplementary Table 4) showed no clear dependence on tau load, and was relatively consistent across cohorts (SSIM_avg_=0.76 to SSIM_avg_=0.85, Supplementary Table 5). Voxel-wise errors differed slightly by diagnosis and tracer, with highest errors in individuals with dementia and in tau-PET regions known for off-target binding (Supplementary Note 1).

#### Synthetic tau-PET reliably captures tau load across multiple brain regions of interest

We next evaluated how accurately tau load (the overall tau burden measured from the PET signal intensity) was estimated in several relevant tau-PET regions of interest (ROIs). Comparing true and synthetic tau-PET Braak ROIs (Fig 3a-b), correlations were R=0.77-0.86 (MRI, age & plasma) or R=0.73-0.80 (MRI & age), while errors were MAE=0.092-0.15 SUVR (MRI, age & plasma) or MAE=0.10-0.16 SUVR (MRI & age). Synthetic tau-PET generated from MRI, age and plasma p-tau217 had significantly higher correlation with tau burden in all Braak ROIs than plasma p-tau217 alone (ΔR=0.059-0.11, p_FDR_ ≤ 0.04, Supplementary Table 4). Across cortical FreeSurfer regions, correlations ranged from R=0.50-0.88 (MRI, age & plasma) or R=0.50-0.85 (MRI & age), with highest performance in regions showing more tau signal on average. Without plasma p-tau217 performance was slightly lower, and the model more often underestimated than overestimated tau load (Fig 3). Results were similar when evaluating the MRI and age input on the subset of participants that had available p-tau217 measures (Supplementary Table 4). Next, we compared errors between diagnostic groups, reporting mean absolute percentage error (MAPE) instead of MAE to account for the between-group differences in tau burden. Highest MAPE was seen for the non-AD dementia group (e.g., Braak I-IV, MAPE=21% versus MAPE=6.2%-15% in the other groups with MRI, age and p-tau217 as input; Supplementary Fig. 2a-c). Errors generally decreased nominally across all diagnostic groups with plasma p-tau217 included as input compared to without (statistically significant only in the NC and SCD groups for Braak I-II load).

**Figure 3:**
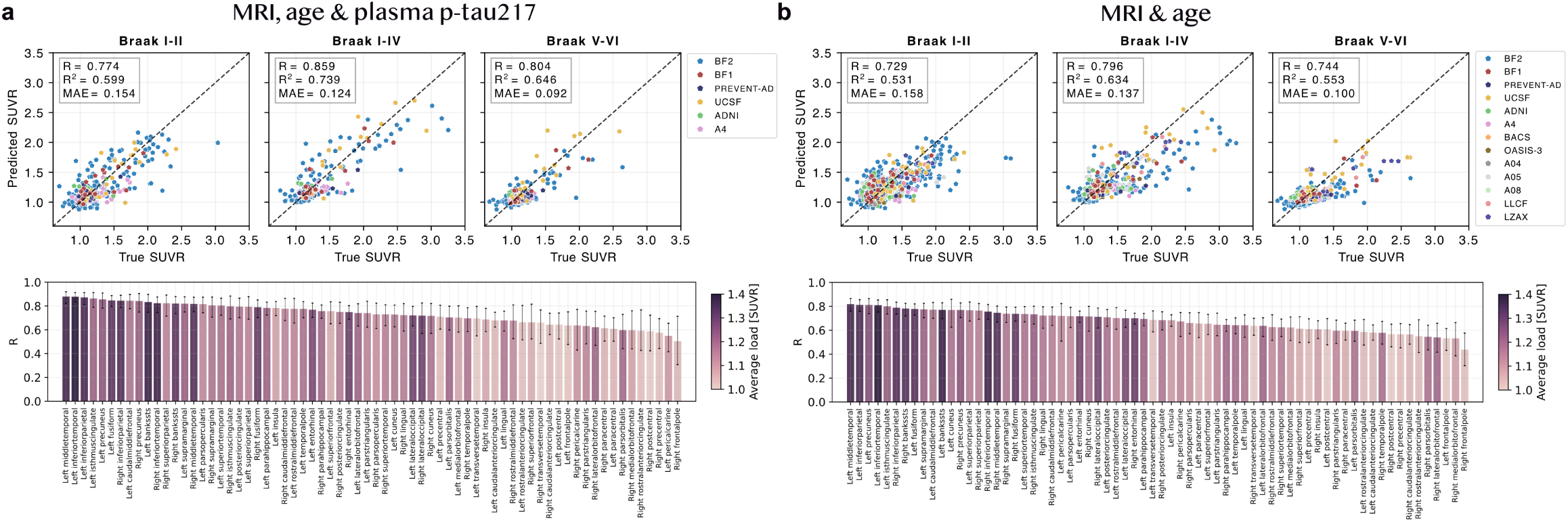
Evaluation of tau load in synthetic compared to true tau-PET in the test set. Scatter plots of true versus predicted (estimated from synthetic) tau-PET load in Braak and cortical FreeSurfer regions when a) using MRI, age and plasma p-tau217 and b) using MRI and age as input to the deep learning model. Abbreviations: Standardized uptake value ratio (SUVR), mean absolute error (MAE).

#### Synthetic tau-PET can model individual tau distribution patterns

Next, the model’s ability to capture spatial patterns of tau deposition was assessed. Within-subject correlations across FreeSurfer regions between synthetic and true PET were on average (std) R_avg_=0.75 (R_std_=0.16) when plasma p-tau217 was included, and R_avg_=0.72 (R_std_=0.16) without (Fig. 4). Variability between subjects was relatively high across all cohorts. Lowest correlations typically occurred in individuals with low overall tau signal, except for one outlier AD dementia case (Supplementary Fig. 3 and Supplementary Table 6, case 19; described in detail in the next section). Restricting the analysis to the subset of individuals with plasma p-tau217 available produced results consistent with those from the full test set sample (Supplementary Table 4). When stratifying by diagnostic group, highest performance was seen for the SCD, MCI and AD dementia groups (e.g., average correlation of R_avg_=0.77 to R_avg_=0.82 [MRI, age and p-tau217]), and lowest in the non-AD dementia group (R_avg_=0.64; Supplementary Fig. 2d). Average correlations increased (statistically significant) for the NC, SCD and other ND groups when including plasma p-tau217 compared to without. We also compared hemispheric differences within the PET images by computing laterality indices across FreeSurfer cortical regions (Fig 4). Mean (std) laterality index correlations were R_avg_=0.56 (R_std_=0.14; MRI, age & p-tau217) or R_avg_=0.48 (R_std_=0.12; MRI & age). Regions with greater average tau asymmetry generally showed higher correlations, likely due to having a larger dynamic range.

**Figure 4:**
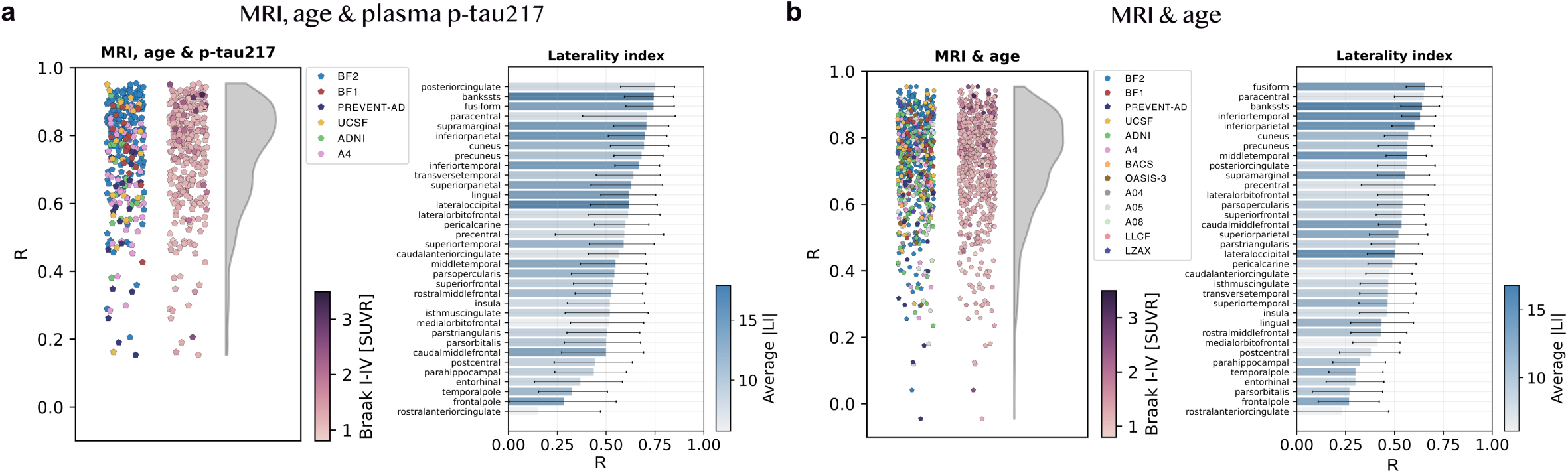
Evaluation of spatial similarity in synthetic compared to true tau-PET in the test set. Scatter plots where each point represents the within-subject regional correlation (based on FreeSurfer segmented volumes) between a participant’s synthetic and true tau-PET. Note that the two columns of scatter points within a subfigure are depicting the same data points only colored differently (by cohort or Braak I-IV load). Each bar in the bar plots represents the correlation across participants between true and synthetic laterality index calculated for each brain region. a) MRI, age and plasma p-tau217 and b) MRI and age as input to the deep learning model. Abbreviations: Standardized uptake value ratio (SUVR), laterality index (LI).

#### Qualitative example cases highlight strengths and limitations of synthetic tau-PET generation

To complement the quantitative evaluation, ten representative cases with varying Braak I-IV burden from multiple cohorts are shown in Fig. 5 (coronal view), Supplementary Fig. 4 (axial and sagittal views), and as animated images in Supplementary videos 1-10. Visually, synthetic images generally appeared to be slightly smoother than true scans. Consistent with the quantitative results, including plasma p-tau217 resulted in more accurate estimation of tau load but had less impact on spatial tau-PET distribution. Performance metrics and basic demographic details for each case are listed in Supplementary Table 6.

**Figure 5:**
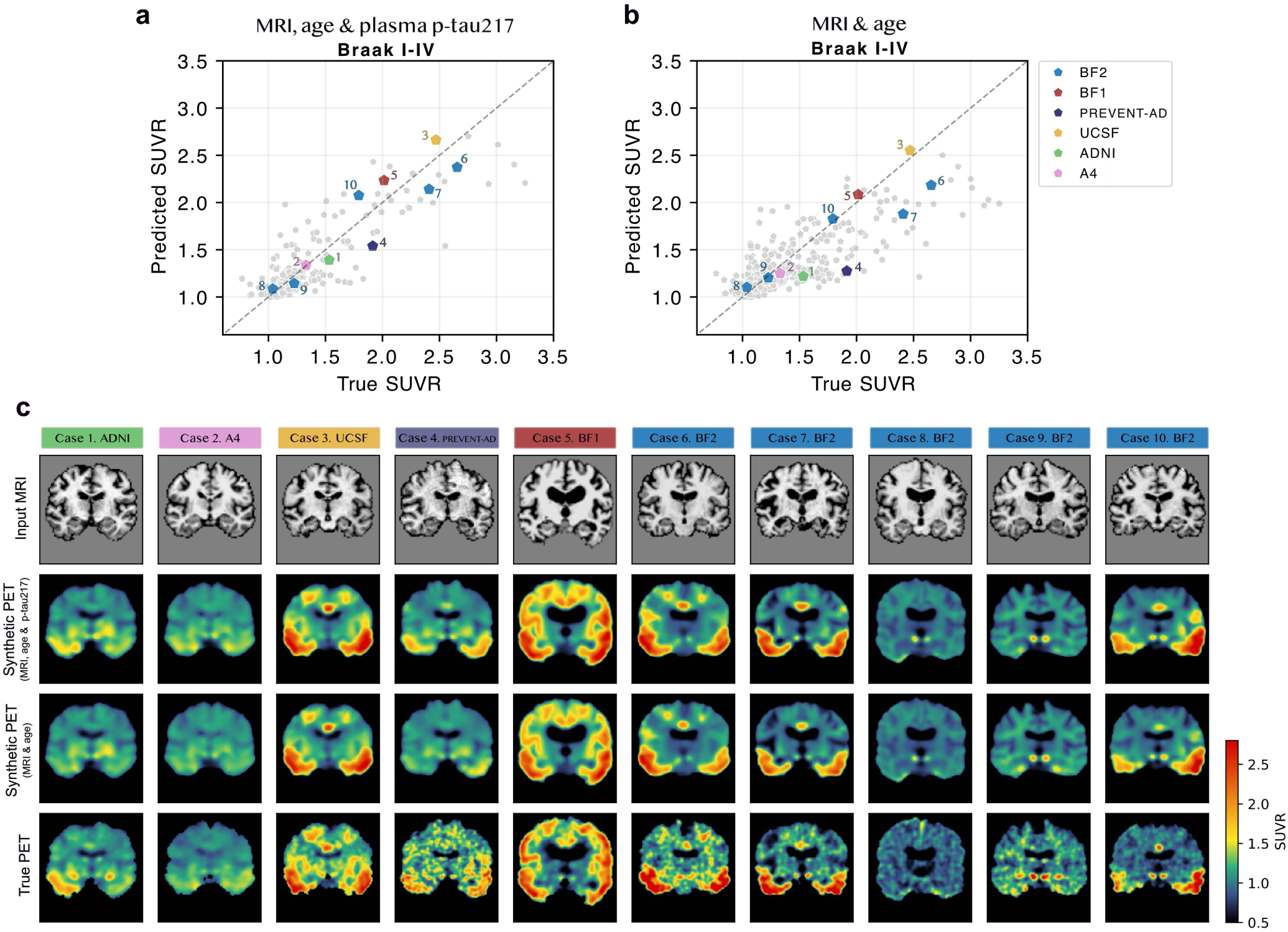
Ten representative example cases. Ten example cases from all cohorts with plasma p-tau217 available. Corresponding Braak I-IV prediction is marked out in a) and b). In c) the input MRI (first row), synthetic tau-PET based on MRI, age and plasma p-tau217 (second row), synthetic tau-PET based on MRI and age (third row), and true tau-PET (fourth row). See Supplementary Fig. 4 for the same cases from axial and sagittal views, and as animated images in Supplementary videos 1-10.

To better understand model strengths and limitations, we examined 20 additional cases: low- and high-performing examples shown in Supplementary Figs. 3 and 5. Cases were selected based on high/low Braak I-IV SUVR error or spatial correlation using visually defined thresholds based on error or correlation distributions, Supplementary Fig. 6. This resulted in low-performing cases of Braak I-IV MAE > 0.60 SUVR or spatial correlation < 0.40, and high-performing of Braak I-IV MAE < 0.10 SUVR or spatial correlation > 0.90. With this definition, low-performing cases were rarer (5.6% of test set) than high (70%; Supplementary Fig. 6). Ten low-performing cases, five with large Braak I–IV errors (cases 11-15) and five with low spatial correlation (cases 16-20), are provided in Supplementary Fig. 3 and Supplementary Table 6. Four high-error cases resulted from underestimation in Aβ-positive AD/MCI individuals (case 11, 12, 14,15), where three of these had outstandingly high true Braak I-IV tau burden (≥2.93 SUVR). Case 13 showed SUVR overestimation for an Aβ-positive behavioral variant frontotemporal dementia (bvFTD) participant that had elevated plasma p-tau217 (0.45 pg/ml) levels in addition to regional atrophy. Interestingly, the six other bvFTD cases in the test set (both Aβ-positives and negatives) showed performance close to the test set average but had lower p-tau217 (≤0.23 pg/ml), likely explaining case 13’s overestimation. Among the low spatial correlation cases, two were AD participants with underestimated medial temporal or frontal signal (cases 17 and 19). The other three (cases 16, 18 and 20) were cognitively unimpaired participants from the PREVENT-AD cohort whose true tau-PET showed atypically diffuse signal, likely reflecting the High-Resolution Research Tomograph (HRRT) used for tau-PET acquisition in this cohort.

Similarly, five cases with very low Braak I-IV error (cases 21-25) and high spatial correlation (cases 26-30) are provided in Supplementary Fig. 5. Since high-performing cases were relatively common and many were Aβ-negative controls without measurable tau pathology, we specifically enriched the example cases for individuals with relatively high true Braak I-IV tau load, yielding mainly Aβ-positive individuals with MCI/AD dementia. Finally, seven representative cases from the cohorts without plasma p-tau217 measurements are provided in Supplementary Fig. 7, illustrating that the model with only MRI and age as input achieved comparable qualitative performance also in these cohorts.

### Assessing potential clinical utility of synthetic tau-PET

#### Synthetic tau-PET captures clinically meaningful prognostic information

To move beyond image metrics, we evaluated whether synthetic tau-PET images provided clinically meaningful information. In 2022, Ossenkoppele, Pichet Binette et al. showed that cognitively unimpaired individuals who are both Aβ-PET and tau-PET positive are at increased risk of developing dementia, which is further refined by regional tau positivity: a 6.2-fold higher risk for A+T_MTL_+ and 41-fold higher risk for A+T_Neo_+ versus A-T-.^27^ We imitated this design in 358 cognitively unimpaired participants from the Swedish BioFINDER-1 cohort (Supplementary Table 7) who had never undergone true tau-PET (and thus were not included in the multi-cohort dataset) but had baseline structural MRI, age, and plasma p-tau217, and a mean follow-up of 6.5 years. Using our trained model, we generated synthetic tau-PET and extracted mean SUVR in medial temporal lobe (T_Syn,MTL_) and neocortical (T_Syn,Neo_) composites. Synthetic tau-positivity was defined as >2 SD above the mean in the A-negative group (similar to ^27^). Kaplan-Meier curves highlighted increased risk of progression to dementia with synthetic tau-positivity, highest in the A+T_Syn,Neo_+ group (Fig. 6a). In Cox regression models, the risk was 12-fold higher in A+T_Syn,MTL_+ (95% CI: 5.2 to 28) and 45-fold higher in A+T_Syn,Neo_+ (95% CI: 19 to 110) compared to A-T_Syn_-, in line with the results previously reported for true tau-PET. Note that this analysis was unsuccessful without plasma p-tau217 in the model (no CU individuals were A+T_Syn,Neo_+).

**Figure 6:**
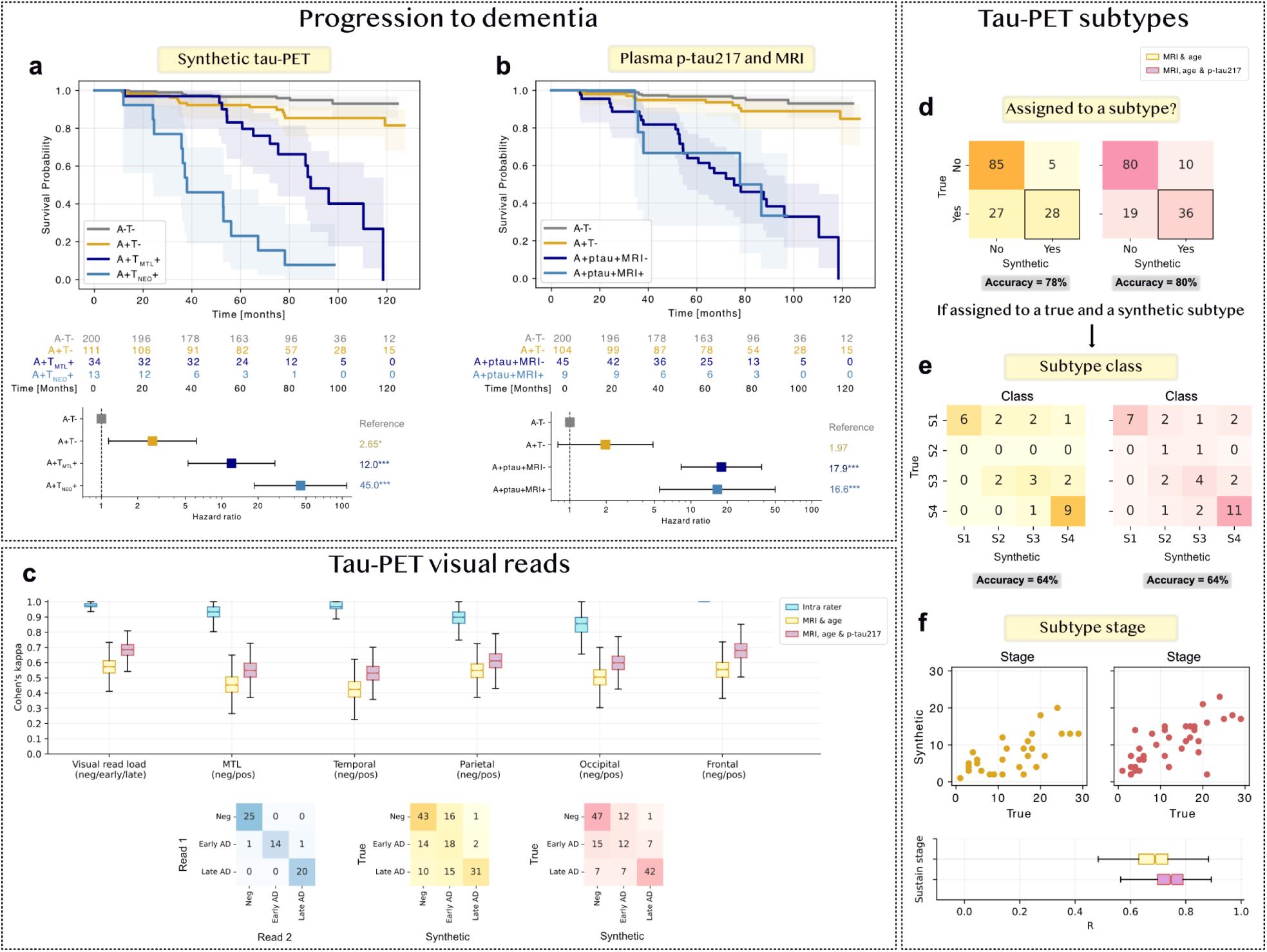
Clinical utility of synthetic tau-PET for prognosis, diagnosis (visual reads), and subtyping. **a)** and b) Kaplan-Meier and Cox regression analyses showing the risk of progression to dementia for cognitively unimpaired individuals in the independent BF1 cohort (participant with no true tau-PET). T-status was derived by a) synthetic tau-PET ROIs or b) plasma p-tau217 and an MRI AD signature ROI. This analysis imitates the study design in Ossenkoppele, Pichet Binette et al. (2022)^27^. c) Expert visual reads of true and synthetic tau-PET for 150 Aβ-positive test-set participants. d), e) and f) Tau-PET Sustain subtyping^9^ directly applied to Aβ-positive test-set participants (n=145), d) assignment to subtype yes or no e) subtype classification and f) subtype stage. Subtype classes: S1 (limbic), S2 (MTL-sparing), S3 (posterior), S4 (lateral temporal). Abbreviations: medial temporal lobe (MTL), Alzheimer’s disease (AD). * p < 0.05, ** p < 0.01, *** p < 0.001

To determine whether the synthetic PET added information beyond what was easily extractable directly from the input data, we compared the prognostic models against a scenario where plasma p-tau217 positivity represented early tau-positivity and MRI atrophy (cortical thickness in an AD-signature composite^30^) represented late tau-positivity. Cutoffs were defined using the same approach (>2 SD above the A-negative mean). This approach also showed increased risk of dementia progression in tau-positive individuals (Fig. 6b), but no clear distinction between early and late tau positivity (A+ptau+MRI-: HR = 17.9; A+ptau+MRI+: HR = 16.6).

#### Synthetic tau-PET show promising but imperfect visual read and subtyping agreement

To evaluate diagnostic utility, an expert rated true and synthetic tau-PET from 150 Aβ-positive test-set participants. Scans were presented in random order without indicating whether they were true or synthetic. The expert classified (1) overall status as negative/inconclusive, early AD, or late AD and (2) positive or negative in five ROIs (medial temporal, temporal, parietal, frontal, and occipital). Cohen’s kappa scores between true and synthetic ratings were 0.69 with plasma p-tau217 and 0.58 without plasma p-tau217 for overall status, and 0.53-0.68 (MRI, age & p-tau217) or 0.42-0.55 (MRI & age) for the five ROIs (Fig. 6c).

To further characterize tau accumulation patterns, we directly applied a fitted SuStaIn tau-PET subtype model^9^ to assess subtype and stage agreement between true and synthetic scans for Aβ-positive test participants. The evaluation sample was limited (A4, BF2 and PREVENT-AD, n=145), as individuals used for SuStaIn training (ADNI, BF1 and UCSF) were excluded to avoid data leakage. Most individuals (62%) were not assigned to a subtype based on true tau-PET. Agreement on subtype assignment (yes or no) between true and synthetic scans was 80% with plasma p-tau217 and 78% without (Fig. 6d). Among participants assigned to a subtype in both the true and synthetic scans (n=36 with plasma p-tau217 and n=28 without), subtype agreement was 64% (Fig. 6e). Stage correlation was R=0.70 with plasma p-tau217 versus R=0.65 without (Fig. 6f).

### Combining plasma p-tau217 and eMTBR-tau243 further enhances model performance

MTBR-tau243 has shown stronger associations with tau-PET burden than p-tau217 in both CSF and plasma.^31,32^ To test whether it could further improve tau-PET synthesis by replacing or complementing plasma p-tau217, we performed exploratory analyses in a BF2 subsample. Although a plasma-based comparison using plasma p-tau217 and eMTBR-tau243 would have been preferable to maintain accessibility, both were available for only n=399 participants. The overlapping plasma p-tau217 and CSF eMTBR-tau243 sample was larger (n=1431; n_train_=1062, n_val_=191, n_test_=178). Given the sample size difference, and since the correlation between CSF eMTBR-tau243 and plasma eMTBR-tau243 was high (R=0.78), we used CSF eMTBR-tau243.

The U-Net using MRI, age, and CSF eMTBR-tau243 performed similarly to the model using MRI, age, and plasma p-tau217 after additional model training (Braak I-IV load R=0.86 versus R=0.85; mean [std] spatial correlation R_avg_=0.80 [R_std_=0.12] versus R_avg_=0.80 [R_std_=0.13]) (Fig. 7a-b). A model incorporating both plasma p-tau217 and CSF eMTBR-tau243 in an additional bottleneck channel performed better than the single-biomarker models (Braak I-IV load R=0.89 and mean [std] spatial correlation R_avg_=0.83 [R_std_=0.095]). Three example scans illustrate cases in which the combined model improved tau load estimates that were slightly over- or underestimated by the single-biomarker models (Fig. 7c; marked out in Fig. 7a, Supplementary Table 8). To test whether replacing or combining plasma p-tau217 with another AD biomarker maintained or improved performance in general, we repeated the analyses with plasma p-tau181. The single p-tau181 model performed similarly to the p-tau217 and eMTBR-tau243 models, but the combined p-tau217 and p-tau181 model performed significantly worse than the combined p-tau217 and eMTBR-tau243 model (Supplementary Table 9).

**Figure 7:**
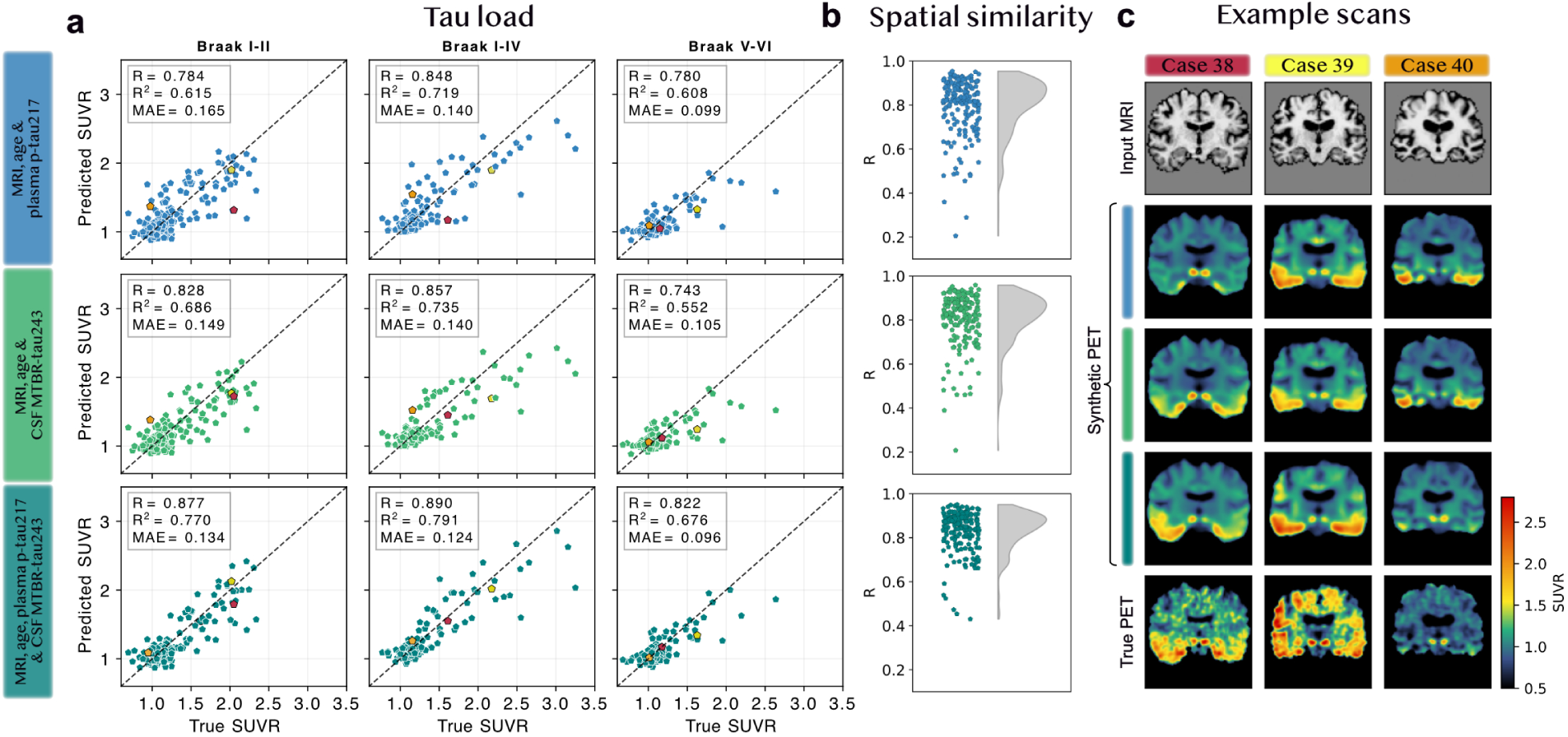
MTBR-tau243 as an alternative or complementary fluid biomarker for tau-PET synthesis. **a)** Tau-PET load estimation of the U-Net using MRI, age, and plasma p-tau217, MRI, age, and CSF eMTBR-tau243, or both plasma p-tau217 and CSF eMTBR-tau243 in the test set with all available data (n=178). **b)** Evaluation of spatial similarity in synthetic compared to true tau-PET in the test set for the same three models (n=178). **c)** Example cases where the combined biomarker model improved tau load estimations (e.g., single biomarker models showed underestimation for Case 38 and 39, and overestimation for Case 40). CSF (and not plasma) eMTBR-tau243 was used for this exploratory analysis to maintain a large sample size (n=1431; n_test_=178).

## Discussion

Our analyses suggest that tau-PET images synthesized from accessible modalities using deep learning closely resembled true images in both quantitative and qualitative assessments. Performance was highest for scans generated with plasma p-tau217 together with MRI and age, but remained strong using only MRI and age, demonstrating flexibility to settings with limited data. In an independent cohort without tau-PET, synthetic scans generated from MRI, age, and plasma p-tau217 predicted progression to dementia in cognitively unimpaired individuals at levels comparable to those reported for true tau-PET^27^, capturing distinct risk profiles associated with early and late tau accumulation. Synthetic images also demonstrated moderate performance for subtyping and expert visual ratings, indicating clinically meaningful signal yet underscoring need for further refinement to reach parity with true tau-PET in these contexts. Preliminary analyses in a subset suggested that combining plasma p-tau217 and eMTBR-tau243 could further boost tau burden estimation. Overall, this study represents a step toward scalable, non-invasive, yet spatially information-rich *in vivo* assessments of tau pathology.

With tau-targeting drugs for AD advancing through clinical development^19,33^, tau-PET will likely become increasingly central for trial recruitment and treatment monitoring. A global survey among dementia experts concluded that tau-PET will likely maintain an important future role in clinical practice and trials.^34^ In this context, tools that approximate tau-PET from more accessible modalities could substantially reduce costs, for example by serving as a pre-screening step to select suitable populations before performing actual tau-PET. Furthermore, the 2025 appropriate use criteria for Aβ-and tau-PET^35^ highlight tau-PET as useful to increase AD diagnostic accuracy and certainty, inform on prognosis, and determine eligibility of Aβ-targeting therapies.^6,36^ Our synthesized scans accurately captured pathological tau load and spatial distribution and predicted progression to dementia in cognitively unimpaired individuals, highlighting the potential usefulness in line with these criteria. Although full replacement of tau-PET is premature, it is important to recognize that tau-PET is costly, requires access to radiotracers, involves radiation, and remains unavailable in most hospitals and memory clinics worldwide.^20^ Thus, non-invasive synthetic tau-PET estimates, treated as approximations, could still provide added diagnostic or prognostic value beyond currently available modalities. Future studies should prospectively assess the clinical utility of synthetic tau-PET to better understand benefits in real-world diagnostic workflows.

In this study, using a fluid biomarker highly predictive of tau pathology like plasma p-tau217^29,37–39^ refined the amplitude of the tau-PET signal inferred via structural MRI. Adding age further improved performance, likely because younger individuals show greater atrophy resilience despite pathological tau^40^, helping the model calibrate tau-related atrophy by age. In contrast, adding sex did not enhance performance, despite reported sex differences in tau burden.^41,42^ This may be due to subtler effects of sex than of age, less informational variability because sex is a binary variable, or that sufficient information about sex already is extractable from the brain MRI.^43^

We systematically tested multiple inputs and U-Net architectural designs, yielding a final model with high performance and robust generalizability. Increasing network depth and filter count consistently improved performance, likely by enhancing representational capacity and expanding the receptive field, allowing each voxel to integrate information from larger spatial contexts. Incorporating residual units further stabilized training and improved performance, while an attention unit in the bottleneck helped the model focus on informative brain regions.^44,45^ Generally, U-Net architectures are widely used and perform strongly across numerous medical imaging tasks, for example image synthesis, super-resolution, and segmentation.^46–49^ However, other architectures, including GANs and diffusion models, also show strong promise for medical image generation and may further improve tau-PET synthesis.^50–54^

There may also exist an upper limit on achievable performance determined by the information contained in the input features, regardless of model architecture. Because MRI-based atrophy occurs downstream of tau accumulation^4^, this time delay may limit the precision with which tau-PET patterns can be recovered from structural MRI. Even when plasma biomarkers are used to calibrate overall tau burden, spatial localization may remain uncertain, particularly in early AD with atypical tau accumulation. Moreover, although plasma p-tau217 is highly predictive of tau pathology, it starts to increase already during Aβ buildup^55–57^, potentially limiting its capacity to fully represent tau load. Our preliminary analyses with eMTBR-tau243 suggest that adding a more tau-specific marker could further refine model performance. Similarly, adding complementary MRI contrasts or other neuroimaging modalities might enhance the model’s ability to synthesize spatial distribution patterns.^58,59^ However, each added modality reduces clinical scalability by increasing complexity and cost per synthetic scan. This creates an inherent trade-off between maximizing performance and maintaining clinical feasibility and accessibility. Ultimately, this trade-off is determined by the requirements and resources of the specific application. In this study, we prioritized simplicity and scalability by focusing on a few widely available input modalities. Future work could more systematically explore this trade-off when adding modalities, to identify an optimal balance between performance and real-world applicability.

Although we included a broad range of diagnoses, non-AD dementias and other neurological disorders were less prevalent and more diverse than AD neuropathological changes. Reasonably, performance was lowest for non-AD dementia participants regarding tau load and spatial distribution patterns. Notably, one low-performing example case with overestimated tau (case 13) was a bvFTD participant with comorbid AD pathology (Aβ-positive and high plasma p-tau217). This case gives insights into model limitations related to comorbidities, specifically highlighting the challenge of distinguishing the driving pathology when multiple exist. If integrating AI tools in clinical use, it will be crucial to recognize such limitations and to integrate additional information, for example clinical syndrome and cognitive profile, to make well-informed decisions.

A visible difference between the true and synthetic tau-PET was smoother synthetic images, likely reflecting the model’s attempt to generate tracer-, scanner- and protocol-independent representations in this heterogeneous multi-cohort setting. As shown by average voxel-wise reconstruction bias (Supplementary note 1), different tracers produced distinct error patterns, partly reflecting known off-target binding characteristics and affinities.^13,60–62^ With a large diverse dataset well-represented by tracers across diagnostic groups, tracer identity could be included as an input, enabling tracer-specific synthetic images. This might disentangle tracer-dependent effects from the common pathological tau signal and thereby improve modeling of true tau. In this study, given additional cohort-specific scanner and acquisition differences and use of two relatively similar tracers ([^18^F]flortaucipir, [^18^F]RO948), we did not pursue this, but it remains an interesting direction for future work.

One limitation with this study was the multi-cohort design. Although a large dataset was needed to train generalizable deep learning models, cohort-related signals (e.g., tracer and scanner differences, partially mitigated by smoothing during pre-processing) limited how precise synthetic scans could align with true ones. However, this also increased the robustness to such variations, potentially beneficial when applying the model in diverse settings. Second, synthetic scans were smoother than most true scans, making real versus synthetic relatively easy to distinguish. Visual reads were hence not fully blinded despite not informing the rater of scan type. One way to mitigate smoothness in future studies could be to incorporate a discriminator into the U-Net architecture (i.e., a GAN framework), to promote generation of more realistic, high-frequency detail. However, this approach risks promoting artificial, non-biological patterns (e.g., tracer-like off-target binding or other “hallucinated” signals) that could compromise interpretability and clinical reliability. Other limitations include the relatively small sample size by deep learning standards, the lack of plasma p-tau217 in several cohorts, and demographic biases (e.g., highly educated participants from Western countries).

In conclusion, we demonstrate that aggregated tau load and spatial distribution measured by PET can be synthesized using more accessible variables in deep learning models. The generated scans informed on both diagnostic and prognostic details, and the model can likely be further improved with larger datasets and complementary input features (e.g., more tau-specific biomarkers or additional MRI modalities). Future studies are needed to establish the effect synthetic tau-PET can have in clinical practice. These findings represent a promising step toward scalable, non-invasive, and spatially detailed in vivo assessment of tau pathology.

## Methods

### Participants

The multi-cohort dataset included 5,191 participants from 13 different AD study cohorts, all previously described in detail: the Anti-Amyloid Treatment in Asymptomatic AD cohort (A4)^63,64^, the Alzheimer’s Disease Neuroimaging Initiative cohort (ADNI; http://adni.loni.usc.edu), the 18F-AV-1451-A04 (A04; NCT02051764), 18F-AV-1451-A05 (A05; NCT02016560), and 18F-AV-1451-A08 (A08; NCT04468347) flortaucipir clinical trials ^65,66^, the I7X-MC-LLCF (NAVIGATE-AD) phase 2 clinical trial (LLCF; NCT02791191)^67^, the H8A-MC-LZAX (EXPEDITION3) phase 3 clinical trial (LZAX; NCT01900665)^68^, the Berkeley Aging Cohort Study (BACS)^69^, a subset of the BioFINDER-1 cohort with tau-PET available (BF1)^38^, the BioFINDER-2 cohort (BF2)^70^, the family history-positive Pre-symptomatic Evaluation of Experimental or Novel Treatments for Alzheimer Disease cohort (PREVENT-AD)^71^, the Open Access Series of Imaging Studies phase 3 cohort (OASIS)^72^, and University of California San Francisco Alzheimer’s Disease Research Center cohort (UCSF)^73,74^. Participants represented a broad spectrum of clinical and cognitive neurodegenerative disorders, including cases with normal cognition (NC, 47.4%), subjective cognitive decline (SCD, 7.53%), mild cognitive impairment (MCI, 19.0%), AD dementia (15.2%), non-AD dementia (4.71%) and other neurological disorders (6.18%). Αβ-status was determined using either Αβ-PET (all cohorts, cutoff 1.26 SUVR in PREVENT-AD; 20 centiloids in all other cohorts)^75^ or CSF Αβ42/Αβ40 (BF1 and BF2, cutoffs 0.066 and 0.08 respectively due to differences in preanalytical handling)^76,77^. All participants had basic demographic information available, and had undergone structural MRI and tau-PET. Plasma p-tau217 was available in a subset of participants (A4, ADNI, BF1, BF2, PREVENT-AD, and UCSF; n=2352). Detailed demographics are presented in Table 1 (by train, validation and test sets) and Supplementary Table 1 (by cohort).

An additional cohort was included to evaluate model performance: 358 cognitively unimpaired BF1 participants that had not undergone tau-PET^78^. The BF1 participants were included in the progression to dementia analysis (average follow-up 6.5 years; Supplementary Table 7).

### Ethics

All participants gave written informed consent to participate, and the studies were approved by local institutional ethical review boards.

### Neuroimaging

#### Image acquisition and preprocessing

All participants underwent 3.0 T MRI acquiring isometric 1 mm^3^ T1-weighted images. Tau-PET was performed using either [^18^F]RO948 (BF2) or [^18^F]flortaucipir (all other cohorts). Standardized uptake value ratio (SUVR) images were created for the 70-90 min ([^18^F]RO948), 75-105 min ([^18^F]flortaucipir; ADNI, Avid cohorts) and 80-100 min ([^18^F]flortaucipir; all other cohorts) post-injection interval using inferior cerebellar cortex as reference regions. All images were centrally processed at Lund University. In brief, each T1-weigthed MRI was skull-stripped and processed using FreeSurfer v.6.0 (https://surfer.nmr.mgh.harvard.edu/) for tissue segmentation and cortical parcellation into ROIs based on the Desikan-Killiany atlas. Tau-PET data were attenuation-corrected, motion-corrected, summed and rigidly co-registered to each participant’s T1-weighted MRI. For each MRI and tau-PET, brain extraction was improved using the FreeSurfer derived brain mask. Each MRI was then rigidly co-registered to Montreal Neurological Institute (MNI) 2×2×2 mm^2^ template space using Advanced Normalization Tools (ANTs) v.2.3.1. The resulting transformation matrix was also applied to the corresponding FreeSurfer-derived masks and tau-PET to ensure spatial alignment across modalities. The FreeSurfer segmentations and parcellations were used to extract mean tau-PET SUVR values within ROIs for both the true and synthetic tau-PET for subsequent regional analyses. Definitions of imaging composites (Braak regions, MRI AD signature, T_MTL_ and T_NEO_)^16,27,30^ are provided in Supplementary Table 10. Tau positivity was defined as Braak I-IV SUVR > 1.36 for [^18^F]RO948 and > 1.34 for [^18^F]flortaucipir.^13,79^

#### Automatic quality control

Before including participants in the multi-cohort dataset, we performed automated quality control procedures to the imaging data to identify and exclude scans with evident technical flaws or misalignments.

1. **Empty tau-PET scans:** We excluded scans that were empty, identified as images where the minimum and maximum voxel intensities were identical (n = 4).
2. **Incomplete FreeSurfer segmentation:** We removed cases missing one or more FreeSurfer regions included in the Braak I–IV composite (n = 5). The missing regions of such scans included the parahippocampal, caudal anterior cingulate, and entorhinal cortices.

#### MRI standard scaling

To account for differences in MRI intensities arising from variations in scanners and acquisition protocols across sites, all MRI scans were intensity-normalized prior to model training. For each individual MRI, we applied voxel-wise standardization (z-scoring) based on the mean and standard deviation of all voxels within the masked brain, ensuring that all images had zero mean and unit variance within the brain. The background was set to zero to help the model easily identify non-brain regions across scans.

#### Tau-PET smoothing

To account for differences in spatial tau-PET resolution across cohorts, we applied smoothing to harmonize the images. For each PET scan, the intrinsic smoothness was first estimated as the full width at half maximum (FWHM) using the AFNI tool 3dFWHMx.^80^ Based on the distribution of estimated FWHM values across all images, we selected a target smoothness corresponding to the 95th upper percentile (FWHM = 7 mm). All PET images with lower intrinsic smoothness were then smoothed using an isotropic Gaussian kernel to reach this level. The harmonized images were used in all analyses.

### Plasma and CSF collection and analysis

Plasma samples were collected in A4, ADNI, BF1, BF2, PREVENT-AD and UCSF and handled according to established preanalytical protocols, previously described in detail.^56,57,64,70,71,74^ Plasma p-tau217 was measured with Eli Lilly immunoassays on a Meso Scale Discovery platform in A4, BF1, BF2, PREVENT-AD and UCSF. Plasma p-tau217 was measured with Fujirebio Lumipulse immunoassay in ADNI, which we bridged to the Lilly assay using n = 1374 in-house samples. CSF eMTBR-tau243 was measured in BF2 with C_2_N Diagnostics mass spectrometry assays. All fluid biomarker analyses were performed by technicians blinded to clinical and imaging data.

### Model development and training

#### U-Net architecture

The U-Net architecture was originally developed by Ronneberger et al.^81^ A U-Net consists of two main components: an encoder that progressively downsamples the input to extract hierarchical features, and a decoder that upsamples these features to reconstruct the output at the original spatial resolution (Figure 2). Both the encoder and decoder are composed of multiple blocks that include convolutional layers (filters), with the number of blocks determining the network’s depth. The encoder and decoder are connected through a bottleneck, representing the most compact, high-level feature representation of the input. A key part of the U-Net architecture is the use of skip connections, which copy the output from each encoder block directly to the corresponding decoder block. These connections allow the network to preserve high-resolution spatial information from the input, while focusing the latent feature space on learning the most relevant differences between input and output data. For example, in the context of generating synthetic tau-PET from MRI, the U-Net design allows structural, anatomical information about the brain to propagate through the skip connections, while biologically meaningful transformations between imaging modalities can be captured through the bottleneck.

#### U-Net blocks

We built our U-Net encoder, bottleneck, and decoder blocks using three main layer compositions: convolutional, residual, and attention units. The convolutional unit formed the basic component of feature extraction, with each such unit consisting of a 3D convolutional layer (kernel size 3×3×3, stride 1) followed by batch normalization (to standardize layer inputs within each batch and thereby reduce internal covariate shift^82^) and a LeakyReLU activation function (to introduce non-linearity while preserving small negative signals to reduce the risk of neurons becoming inactive^83^). The residual unit extended the convolutional unit by introducing an additional 3D convolution layer (kernel size 3×3×3, stride 1) with batch normalization, combined through a residual connection. The residual connection improves training stability for deeper networks by providing a shortcut path for the gradient to propagate, which reduces the risk of vanishing gradients.^44^ LeakyReLU was again used as the activation function. Finally, the attention unit, which we based on query-key-value formulation,^45^ learns to highlight the most relevant spatial features by computing attention weights and reweighting the input features accordingly. Through this mechanism, the network can selectively emphasize regions of more interest.^84^ For downsampling in the encoder we used max pooling, and for upsampling in the decoder transposed convolutions. An overview of the arrangement of these units within the encoder-decoder blocks of our final model is shown in Figure 2, with implementation details in https://github.com/DeMONLab-BioFINDER/karlsson_synthetic_taupet.

#### Loss function

We defined a loss function ℒ that was an L1-based combination of two terms. The first term was a standard L1 loss, measuring the average absolute difference between the true and predicted image *x*, *x^* ∈ ℝ^H×W×D^ across all N*_v_*voxels. The second term was a masked L1 loss, where the error was computed only in voxels selected by a predefined mask *m* ∈ {0,1}^H×W×D^ (e.g., brain regions of interest) and scaled by a scalar weight α:

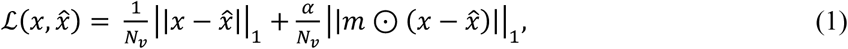

where ⊙ denotes elementwise multiplication. This formulation encourages the model to minimize global reconstruction error while paying extra attention to (pre-defined) biologically relevant regions.

In our study, we set α = ½ and used the FreeSurfer cortical mask as *m* to make the model slightly more focused on cortical gray matter.

#### Training

Model optimization was performed using the Adaptive Moment Estimation (Adam) optimizer^85^, which adapts learning rates for individual parameters based on estimates of first- and second-order moments of the gradients. The Adam optimizer is particularly useful for stabilizing training with small batch sizes and variable gradient magnitudes. The model was trained in two stages. We started with 120 epochs using random initialization and a learning rate of 0.0001 to enable broad exploration of the optimization landscape and escape shallow minima. This was followed by fine-tuning from the identified minimum using a lower learning rate (1×10^-6^ or 1×10^-7^) to refine convergence. The model weights corresponding to the minimum validation loss were selected.

#### Systematic model development

To design a high-performing U-Net for generating synthetic tau-PET from MRI and other accessible data, we systematically evaluated alternative model designs and input data configurations. This process included standard hyperparameter tuning (e.g., learning rate, batch size, dropout rate, and kernel size) as well as systematic experimentation to understand the contribution of different input features, how to efficiently integrate tabular variables, and to find a suitable network architecture. Key decisions and findings are summarized below, reflecting the main take-aways of the comprehensive and iterative model development process.

#### Input feature configuration

To systematically evaluate the contribution from different input features, we first trained a compact 3D U-Net (∼6M trainable parameters, depth=4, number of filters=16-256) exploring several input configurations. This small-scale network allowed efficient exploration of feature configurations at a low computational cost. The initial model for benchmarking used only MRI as input. Additional variables p-tau217, age and sex were incorporated one at a time to evaluate their individual contributions. Each variable was inputted as an additional channel to the bottleneck. Missing plasma p-tau217 values were imputed to the training set mean to ensure that participants without plasma data could still be included during training. Performance gains were observed when adding age or plasma p-tau217, whereas sex did not contribute to a distinct improvement (Supplementary Table 2). We therefore combined MRI, age, and plasma p-tau217 into a joint model, further improving performance.

#### Integration of tabular variables

We next investigated alternative strategies of integrating tabular variables within the network. To ensure that all spatial locations in the image could interact with the tabular information, each variable was incorporated as a full additional channel at different locations within the network. We hypothesized that tabular variables would be most effectively incorporated at the U-Net bottleneck, as this latent feature space represents the most compact and informative representation of the input. In contrast, including tabular variables at the input layer could lead to attenuation of this information, as it would need to propagate through the entire encoder before influencing the latent features. We tested this hypothesis and observed that the loss function converged to a lower value when tabular variables were added at the bottleneck than to the first encoder block, Supplementary Table 2.

#### Model architecture and size

After selecting input configuration (MRI, age and plasma p-tau217) and integration of tabular variables (bottleneck), we scaled the architecture to improve representational capacity. This included increasing the number of convolutional filters and encoder-decoder blocks (depth), as well as systematically evaluating alternative block designs (e.g. adding or removing convolutional, residual or attention units in encoder/decoder blocks or bottleneck). For each design, we also tuned hyperparameters (e.g., learning rate, batch size, dropout rate, and kernel size). We report the lowest validation loss achieved for each architecture in Supplementary Table 2. The best-performing and final model was a 3D U-Net with ∼110M trainable parameters. The encoder-decoder blocks included convolution and residual units, while the bottleneck included convolutional and attention units (Figure 2).

#### Adding CSF MTBR-tau243 as an additional input

When training the model with the additional fluid biomarker, the same U-Net architecture as in Figure 2 was used but with one additional input channel in the bottleneck. The whole network was re-trained from scratch with the full training and validation sets. The same strategy as for plasma p-tau217 was applied to participants missing CSF MTBR-tau243 values: they were imputed to the training set mean to ensure that participants without CSF MTBR-tau243 data could still be included during training.

### Visual reads

Visual reads of scans from Aβ-positive participants in the test set were performed by an expert rater. In total, 650 scans were evaluated: 150 × 3 (true scans, synthetic scans generated with plasma p-tau217, and synthetic scans generated without plasma p-tau217), 75 × 2 (true scans and synthetic scans generated without plasma p-tau217), and 50 true tau-PET duplicates used to assess intra-rater variability. Before rating, the expert reviewed 20 practice cases in which the Braak I–II, Braak I–IV, and Braak V–VI SUVR levels were displayed, to calibrate the expert’s ratings to this context (e.g., color scale and skull-stripped brain). These 20 practice cases were true scans from the validation set, enriched for cases with Braak I–IV signal around 1.3 SUVR (typically considered borderline positive and challenging). The 650 scans were then randomly shuffled, and no information whether a scan was true or synthetic was given to the rater. The expert classified scans as negative/inconclusive, early AD, or late AD, and additionally rated them as positive or negative in five regions: medial temporal lobe, temporal, parietal, frontal, and occipital cortices.

### Statistical analyses

Analyses were performed using Python v.3.9 or R v.4.2. The final model was evaluated on unseen test data. Full image quality was assessed using mean absolute error (MAE) and structural similarity index measure (SSIM) between true and synthetic scans. Tau load in ROIs of true and synthetic scans were compared using Pearson correlation coefficient (R), proportion of explained variance (R^2^), mean absolute error (MAE) and mean absolute percentage error (MAPE). Within-subject correlations across FreeSurfer cortical regions were calculated by extracting the mean signal in each region of true and synthetic scans and computing the Pearson correlation coefficient. Laterality index (LI) was calculated as the difference between the left and right hemispheres normalized to the average signal in the left and right hemispheres:^29,86^

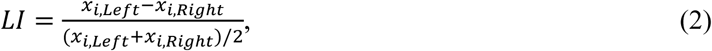

where *x*_i_ is the SUVR of brain region *i*. Average error maps were computed by voxel-wise subtraction of true PET from synthetic PET and averaging across groups. Cox proportional-hazards models were used to calculate hazard ratios for progression to dementia. Thresholds of T positivity were defined as >2 SD above the mean of Αβ-negative participants. Visual ratings were evaluated on Cohen’s kappa with quadratic weighting. Tau-PET subtype classification for true and synthetic scans was performed by applying the fitted Sustain model from Vogel et al. (2021)^9^ to Aβ-positive participants in the test set. Individuals that were used to train the original SuStaIn model (including ADNI, BF1 and UCSF) were excluded to avoid data leakage. During performance comparisons stratified by clinical diagnostic group, two-tailed significance testing was performed as bootstrapped difference between input configurations for a given metric. A p-value <0.05 after adjusting for multiple comparisons by the Benjamini–Hochberg false discovery rate (FDR) method was considered statistically significant.

### Computational resources

Computations were performed on the Bianca Cluster, which is a part the National Academic Infrastructure for Supercomputing in Sweden (NAISS). Bianca is dedicated to analyses of sensitive data, providing 4480 cores in the form of 204 dual CPU (Intel Xeon E5-2630 v3) Huawei XH620 V3 nodes with 128 GB memory, and ten nodes with two NVIDIA A100 40GB GPUs each.

## Code availability

The code and model will be made available at https://github.com/DeMONLab-BioFINDER/karlsson_synthetic_taupet.

## Data Availability

Participant data from open access cohorts (ADNI, A4, OASIS and PREVENT-AD) can be obtained from http://adni.loni.usc.edu/, https://ida.loni.usc.edu/, https://sites.wustl.edu/oasisbrains/, and https://openpreventad.loris.ca/. Pseudonymized data from the other cohorts (BF1, BF2, Avid studies, BACS, UCSF) can be shared with qualified academic researcher upon request to respective principal investigator (for UCSF: submit a request form at https://memory.ucsf.edu/research-trials/professional/open-science) for the purpose of replicating procedures and results presented in the study. Data transfer must be in agreement with the data protection regulation at the institution and decisions by the local ethics review board.

## Acknowledgements

We thank all participants and staff involved in the A4, ADNI, Avid, BACS, BioFINDER, PREVENT-AD, OASIS-3, and UCSF study groups. Work at the authors’ research center was supported by the National Institute of Aging (grant no. R01AG083740), European Research Council (grant no. ADG-101096455), Alzheimer’s Association (grant nos. ZEN24-1069572 and SG-23-1061717), GHR Foundation, Michael J. Fox Foundation (grant no. MJFF-025507), Lilly Research Award Program, WASP and DDLS Joint call for research projects (grant no. WASP/DDLS22-066), Swedish Research Council (grant nos. 2021-02219, 2022-00775 and 2018-02052), ERA PerMed (grant no. ERAPERMED2021-184), Knut and Alice Wallenberg Foundation (grant no. 2022-0231), Strategic Research Area MultiPark (Multidisciplinary Research in Parkinson’s disease) at Lund University, Swedish Alzheimer Foundation (grant nos. AF-1011949, AF-1012595, AF-994229 and AF-980907), Swedish Brain Foundation (grant nos. FO2023-0163, FO2024-0284 and FO2021-0293), Parkinson foundation of Sweden (grant no. 1412/22), Cure Alzheimer’s fund, Rönström Family Foundation (grant nos. FRS-0003, FRS-0004, FRS-0011 and FRS-0013), Berg Family Foundation, Ingvar Kamprad Foundation (grant no. 20243058), Avid Pharmaceuticals, Bundy Academy, Konung Gustaf V:s och Drottning Victorias Frimurarestiftelse, Skåne University Hospital Foundation (grant no. 2020-O000028), Regionalt Forskningsstöd (grant nos. 2022-1259 and 2022-1346) and Swedish federal government under the ALF agreement (2022-Projekt0080, 2022-Projekt0107). JWV is supported by the SciLifeLab & Wallenberg Data Driven Life Science Program (KAW 2020.0239), the Swedish Brain Foundation (FO2025-0307-HK-247), the Swedish Research Council (2024-03642) and the European Research Council (StG-101221737). Data collection and sharing for this project was funded by the Alzheimer’s Disease Neuroimaging Initiative (ADNI) (National Institutes of Health Grant U01 AG024904) and DOD ADNI (Department of Defense award number W81XWH-12-2-0012). ADNI is funded by the National Institute on Aging, the National Institute of Biomedical Imaging and Bioengineering, and through generous contributions from the following: AbbVie, Alzheimer’s Association; Alzheimer’s Drug Discovery Foundation; Araclon Biotech; BioClinica, Inc.; Biogen; Bristol-Myers Squibb Company; CereSpir, Inc.; Cogstate; Eisai Inc.; Elan Pharmaceuticals, Inc.; Eli Lilly and Company; EuroImmun; F. Hoffmann-La Roche Ltd and its affiliated company Genentech, Inc.; Fujirebio; GE Healthcare; IXICO Ltd.; Janssen Alzheimer Immunotherapy Research & Development, LLC.; Johnson & Johnson Pharmaceutical Research & Development LLC.; Lumosity; Lundbeck; Merck & Co., Inc.; Meso Scale Diagnostics, LLC.; NeuroRx Research; Neurotrack Technologies; Novartis Pharmaceuticals Corporation; Pfizer Inc.; Piramal Imaging; Servier; Takeda Pharmaceutical Company; and Transition Therapeutics. The Canadian Institutes of Health Research is providing funds to support ADNI clinical sites in Canada. Private sector contributions are facilitated by the Foundation for the National Institutes of Health (www.fnih.org). The grantee organization is the Northern California Institute for Research and Education, and the study is coordinated by the Alzheimer’s Therapeutic Research Institute at the University of Southern California. ADNI data are disseminated by the Laboratory for Neuro Imaging at the University of Southern California. The A4 Study was a secondary prevention trial in preclinical Alzheimer’s disease, aiming to slow cognitive decline associated with brain amyloid accumulation in clinically normal older individuals. The A4 Study was funded by a public-private-philanthropic partnership, including funding from the National Institutes of Health-National Institute on Aging, Eli Lilly and Company, Alzheimer’s Association, Accelerating Medicines Partnership, GHR Foundation, an anonymous foundation, and additional private donors, with in-kind support from Avid Radiopharmaceuticals, Cogstate, Albert Einstein College of Medicine and the Foundation for Neurologic Diseases.The companion observational Longitudinal Evaluation of Amyloid Risk and Neurodegeneration (LEARN) Study was funded by the Alzheimer’s Association and GHR Foundation. The A4 and LEARN Studies were led by Dr. Reisa Sperling at Brigham and Women’s Hospital, Harvard Medical School, and Dr. Paul Aisen at the Alzheimer’s Therapeutic Research Institute (ATRI) at the University of Southern California. The A4 and LEARN Studies were coordinated by ATRI at the University of Southern California, and the data are made available under the auspices of Alzheimer’s Clinical Trial Consortium through the Global Research & Imaging Platform (GRIP). The complete A4 Study Team list is available on: https://www.actcinfo.org/a4-study-team-lists/. We would like to acknowledge the dedication of the study participants and their study partners who made the A4 and LEARN Studies possible. The data contributed from the PREVENT-AD was supported by public–private partnership funds provided by McGill University, the Fonds de Recherche du Québec — Santé, an unrestricted research grant from Pfizer Canada, the Levesque Foundation, the Douglas Hospital Research Centre and Foundation, the Government of Canada, the Canada Fund for Innovation, the Canadian Institutes of Health Research, the Alzheimer Society of Canada, the National Institutes of Health of the United States, the Alzheimer Association and Brain Canada Foundation. Data were provided in part by OASIS-3, Principal Investigators: T. Benzinger, J. Morris; NIH P30 AG066444, AW00006993. AV-1451 doses were provided by Avid Radiopharmaceuticals, a wholly owned subsidiary of Eli Lilly. The funding sources had no role in the design and conduct of the study; in the collection, analysis and interpretation of the data; or in the preparation, review or approval of the paper.

