## Supplementary Tables for "Generating synthetic tau-PET scans in Alzheimer’s disease from MRI, blood biomarkers and demographics with deep learning"

**Supplementary Table 1: Participant demographics by cohort.** Characteristics of the multi-cohort dataset by cohorts.

|  | **A4** | **ADNI** | **A04** | **A05** | **A08** | **LLCF** | **LZAX** | **BACS** | **BioFINDER-1** | **BioFINDER-2** | **PREVENT-AD** | **OASIS** | **UCSF** |
| --- | --- | --- | --- | --- | --- | --- | --- | --- | --- | --- | --- | --- | --- |
| **Number of participants** | 446 | 857 | 40 | 245 | 82 | 87 | 103 | 134 | 242 | 2130 | 134 | 440 | 251 |
| **Number of participants with plasma p-tau217** | 357 | 122 | 0 | 0 | 0 | 0 | 0 | 0 | 163 | 1494 | 102 | 0 | 114 |
| **Age, mean (std) years** | 71.8 (4.84) | 71.4 (6.77) | 71.8 (7.91) | 68.6 (14.2) | 75.2 (6.59) | 72.5 (7.38) | 73.3 (8.43) | 77.2 (5.51) | 71.1 (7.88) | 69.1 (11.9) | 67.3 (4.11) | 69.7 (8.00) | 64.3 (8.11) |
| **Sex n (%) female** | 257 (57.6) | 441 (51.5) | 9 (22.5) | 120 (49.0) | 47 (57.3) | 52 (59.8) | 56 (54.4) | 57 (42.5) | 114 (47.1) | 1047 (49.2) | 94 (70.1) | 239 (54.3) | 118 (47.0) |
| **Education level, mean (std) years** | 16.2 (2.83) | 16.4 (2.49) | 15.8 (1.99) | 15.7 (2.47) | 14.4 (3.02) | 14.6 (2.84) | 14.3 (3.29) | 17.1 (2.47) | 12.2 (3.67) | 12.7 (3.72) | 15.3 (3.31) | 16.3 (2.39) | 17.0 (3.32) |
| ***APOE* ε4 alleles*, n (%)** |  |  |  |  |  |  |  |  |  |  |  |  |  |
| 0 | - | 467 (60.3) | - | 155 (63.3) | 77 (93.9) | 24 (27.9) | 31 (31.6) | 81 (74.3) | 85 (45.9) | 1134 (53.8) | 63 (61.8) | 251 (60.3) | 77 (60.2) |
| 1 | - | 247 (31.9) | - | 70 (28.6) | - | 47 (54.7) | 50 (51.0) | 28 (25.7) | 71 (38.4) | 814 (38.6) | 38 (37.3) | 142 (34.1) | 40 (31.2) |
| 2 | - | 60 (7.75) | - | 20 (8.16) | 5 (6.10) | 15 (17.4) | 17 (17.4) | - | 29 (15.7) | 161 (7.63) | 1 (0.98) | 23 (5.53) | 11 (8.59) |
| **Clinical diagnosis*, n (%)** |  |  |  |  |  |  |  |  |  |  |  |  |  |
| NC | 446 (100) | 467 (55.5) | 24 (60.0) | 81 (33.1) | 23 (28.0) | - | - | 134 (100) | 53 (25.4) | 670 (31.9) | 134 (100) | 358 (86.1) | 2 (0.957) |
| SCD | - | - | - | - | 43 (52.4) | - | - | - | 12 (5.74) | 323 (15.4) | - | - | 2 (0.957) |
| MCI | - | 281 (33.4) | 11 (27.5) | 111 (45.3) | 12 (14.6) | - | - | - | 33 (15.8) | 472 (22.5) | - | 3 (0.721) | 34 (16.3) |
| AD dementia | - | 82 (9.75) | 5 (12.5) | 53 (21.6) | 4 (4.88) | 87 (100) | 101 (100) | - | 50 (23.9) | 290 (13.8) | - | 43 (10.3) | 51 (24.4) |
| Non-AD dementia | - | 11 (1.31) | - | - | - | - | - | - | 25 (12.0) | 132 (6.28) | - | 12 (2.88) | 58 (27.8) |
| Other neurological disorders | - | - | - | - | - | - | - | - | 36 (17.2) | 214 (10.2) | - | - | 62 (29.7) |
| **Aβ positive*, n (%)** | 379 (85.0) | 371 (44.0) | 34 (85.0) | 94 (38.4) | 32 (39.0) | 87 (100) | 101 (100) | 45 (41.3) | 106 (75.2) | 973 (47.5) | 22 (21.6) | 144 (34.6) | 98 (54.7) |
| **Tau-PET tracer** | [^18^F]flor-taucipir | [^18^F]flor-taucipir | [^18^F]flor-taucipir | [^18^F]flor-taucipir | [^18^F]flor-taucipir | [^18^F]flor-taucipir | [^18^F]flor-taucipir | [^18^F]flor-taucipir | [^18^F]flor-taucipir | [^18^F]RO948 | [^18^F]flor-taucipir | [^18^F]flor-taucipir | [^18^F]flor-taucipir |
| **Plasma p-tau217*, mean (std) pg/ml** | 0.274 (0.150) | 0.370 (0.277) | - | - | - | - | - | - | 0.614 (2.46) | 0.312 (0.299) | 0.267 (0.232) | - | 0.445 (0.374) |

Non-AD dementia includes Frontotemporal dementia, Lewy body dementia, Vascular dementia, and Posterior cortical atrophy. Other neurological disorder includes Parkinson's disease, Progressive supranuclear palsy, Corticobasal syndrome, multiple system atrophy, traumatic brain injury, chronic traumatic encephalopathy, and bipolar disorder.

Abbreviations: normal cognition (NC), subjective cognitive decline (SCD), mild cognitive impairment (MCI).

*missing data for some participants

**Supplementary Table 2: Systematic model development model designs and input data configurations.**

| **Input features** | **Model params** | | | | | **Loss** | |
| --- | --- | --- | --- | --- | --- | --- | --- |
|  | Depth | # filters | # train. params | Int. tab var. | Encoder/Decoder blocks  (Differences from final model in Figure 2) |  |  |
| *Input feature configuration* | | | | | | | |
| MRI | 4 | min: 16  max: 256 | 6M | Bottleneck | Without residual unit in decoder | | 13.62 |
| MRI, age | 4 | min: 16  max: 256 | 6M | Bottleneck | Without residual unit in decoder | | 13.54 |
| MRI, sex | 4 | min: 16  max: 256 | 6M | Bottleneck | Without residual unit in decoder | | 13.61 |
| MRI, p-tau217 | 4 | min: 16  max: 256 | 6M | Bottleneck | Without residual unit in decoder | | 13.48 |
| *Integration of tabular variables* | | | | | | | |
| MRI, age, p-tau217 | 4 | min: 16  max: 256 | 6M | Bottleneck | Without residual unit in decoder | | 13.43 |
| MRI, age, p-tau217 | 4 | min: 16  max: 256 | 6M | Initial | Without residual unit in decoder | | 13.51 |
| *Model architecture and size* | | | | | | | |
| MRI, age, p-tau217 | 4 | min: 32  max: 512 | 28M | Bottleneck | Without residual unit in decoder | | 13.33 |
| MRI, age, p-tau217 | 5 | min: 16  max: 512 | 28M | Bottleneck | Without residual unit in decoder | | 13.26 |
| MRI, age, p-tau217 | 5 | min: 32  max: 1024 | 92M | Bottleneck | Without residual unit in decoder | | 13.25 |
| MRI, age, p-tau217 | 5 | min: 32  max: 1024 | 91M | Bottleneck | Without residual unit in decoder, without attention unit in bottleneck | | 13.25 |
| MRI, age, p-tau217 | 5 | min: 32  max: 1024 | 100M | Bottleneck | Without residual unit in decoder, with extra convolutional unit in encoder. | | 13.44 |
| MRI, age, p-tau217 | 5 | min: 32  max: 1024 | 120M | Bottleneck | With two convolutional units in encoder. | | 13.37 |
| MRI, age, p-tau217 | 5 | min: 32  max: 1024 | 83M | Bottleneck | Without convolutional unit in encoder, without residual unit in decoder, with attention unit in decoder. | | 13.58 |
| **MRI, age, p-tau217** | **5** | **min: 32**  **max: 1024** | **110M** | **Bottleneck** | **FINAL MODEL (Figure 2).** | | **13.10** |

**Supplementary Table 3: Test set correlation with true tau-PET load in Braak regions comparing synthetic tau-PET generated from MRI, age and plasma p-tau217 with raw plasma p-tau217 values (n=318).**

|  | **Synthetic tau-PET  (MRI, age & p-tau217)**  **R (95% CI)** | **Plasma p-tau217**  **R (95% CI)** | **ΔR** | **p-value**  **(FDR corrected)** |
| --- | --- | --- | --- | --- |
| **Braak I-II** | 0.77 (0.71 – 0.82) | 0.66 (0.58 – 0.74) | 0.11 | **0.0027** |
| **Braak I-IV** | 0.86 (0.81 – 0.90) | 0.77 (0.71 – 0.83) | 0.085 | **0.0027** |
| **Braak V-VI** | 0.80 (0.72 – 0.87) | 0.74 (0.65 – 0.82) | 0.059 | **0.040** |

**Supplementary Table 4: Model performance for all individuals and for tau-positives only.**

|  | MRI, age &  p-tau217 | MRI & age  (subsample) | MRI & age  (full sample) |
| --- | --- | --- | --- |
| All individuals | | | |
| n | 318 | 318 | 678 |
| Mean SSIM | 0.811 | 0.809 | 0.810 |
| Mean MAE full image | 0.075 | 0.075 | 0.074 |
| MAE Braak I-IV (load) | 0.124 | 0.147 | 0.137 |
| R Braak I-IV (load) | 0.859 | 0.781 | 0.796 |
| R^2^ Braak I-IV (load) | 0.739 | 0.610 | 0.634 |
| Mean within-subject R (spatial) | 0.745 | 0.726 | 0.721 |
| Mean within-subject R^2^ (spatial) | 0.579 | 0.554 | 0.544 |
| Tau-positives | | | |
| n | 68 | 68 | 153 |
| Mean SSIM | 0.813 | 0.802 | 0.808 |
| Mean MAE full image | 0.106 | 0.110 | 0.105 |
| MAE Braak I-IV (load) | 0.280 | 0.385 | 0.333 |
| R Braak I-IV (load) | 0.750 | 0.668 | 0.695 |
| R^2^ Braak I-IV (load) | 0.563 | 0.447 | 0.484 |
| Mean within-subject R (spatial) | 0.785 | 0.754 | 0.774 |
| Mean within-subject R^2^ (spatial) | 0.640 | 0.598 | 0.618 |

**Supplementary Table 5: Model performance by cohort.**

|  | MRI, age & p-tau217 | | | | MRI & age | | | |
| --- | --- | --- | --- | --- | --- | --- | --- | --- |
|  | n | Mean SSIM | Mean MAE full image | MAE Braak I-IV | n | Mean SSIM | Mean MAE full image | MAE Braak I-IV |
| A4 | 57 | 0.840 | 0.0567 | 0.0824 | 63 | 0.841 | 0.0565 | 0.0869 |
| ADNI | 20 | 0.857 | 0.0518 | 0.0704 | 105 | 0.841 | 0.0568 | 0.0864 |
| A04 | - | - | - | - | 8 | 0.822 | 0.0560 | 0.0540 |
| A05 | - | - | - | - | 46 | 0.826 | 0.0657 | 0.126 |
| A08 | - | - | - | - | 15 | 0.812 | 0.0600 | 0.0487 |
| LLCF | - | - | - | - | 8 | 0.845 | 0.0861 | 0.179 |
| LZAX | - | - | - | - | 16 | 0.807 | 0.131 | 0.250 |
| BACS | - | - | - | - | 8 | 0.827 | 0.0620 | 0.0597 |
| BioFINDER-1 | 18 | 0.819 | 0.0745 | 0.128 | 23 | 0.819 | 0.0724 | 0.144 |
| BioFINDER-2 | 184 | 0.800 | 0.0804 | 0.140 | 311 | 0.794 | 0.0810 | 0.166 |
| PREVENT-AD | 19 | 0.760 | 0.0780 | 0.0928 | 21 | 0.757 | 0.0786 | 0.0957 |
| OASIS | - | - | - | - | 23 | 0.776 | 0.0686 | 0.0724 |
| UCSF | 20 | 0.829 | 0.0931 | 0.177 | 31 | 0.821 | 0.0944 | 0.221 |

| Example Case | SSIM  (w. p-tau217) | SSIM (age MRI only) | True Braak I-IV load | Synthetic Braak I-IV load (w. p-tau217) | Synthetic Braak I-IV load (age MRI only) | Spatial correlation (w. p-tau217) | Spatial correlation (age MRI only) | Age | Sex | Diagnosis | Aβ status | Cohort |
| --- | --- | --- | --- | --- | --- | --- | --- | --- | --- | --- | --- | --- |
| Representative example cases (Fig. 5, Supplementary Fig. 4) | | | | | | | | | | | | |
| Case 1 | 0.872 | 0.851 | 1.53 | 1.39 | 1.22 | 0.776 | 0.625 | 74 | F | CN | + | ADNI |
| Case 2 | 0.867 | 0.860 | 1.33 | 1.34 | 1.25 | 0.766 | 0.743 | 70 | F | CN | + | A4 |
| Case 3 | 0.876 | 0.881 | 2.47 | 2.66 | 2.55 | 0.951 | 0.954 | 53 | F | ADD | + | UCSF |
| Case 4 | 0.771 | 0.738 | 1.91 | 1.54 | 1.27 | 0.753 | 0.632 | 72 | F | CN | + | PREVENT-AD |
| Case 5 | 0.826 | 0.827 | 2.01 | 2.23 | 2.08 | 0.888 | 0.882 | 64 | M | ADD | + | BF1 |
| Case 6 | 0.790 | 0.789 | 2.66 | 2.37 | 2.18 | 0.889 | 0.894 | 57 | M | ADD | + | BF2 |
| Case 7 | 0.854 | 0.851 | 2.41 | 2.14 | 1.88 | 0.917 | 0.925 | 74 | M | ADD | + | BF2 |
| Case 8 | 0.773 | 0.765 | 1.04 | 1.09 | 1.10 | 0.788 | 0.703 | 70 | M | MCI | - | BF2 |
| Case 9 | 0.838 | 0.838 | 1.23 | 1.14 | 1.20 | 0.814 | 0.846 | 75 | M | MCI | + | BF2 |
| Case 10 | 0.859 | 0.866 | 1.79 | 2.08 | 1.83 | 0.922 | 0.923 | 76 | F | ADD | + | BF2 |
| Low-performing example cases (Supplementary Fig. 3) | | | | | | | | | | | | |
| Case 11 | 0.855 | 0.823 | 2.93 | 2.20 | 1.87 | 0.912 | 0.933 | 49 | F | ADD | + | UCSF |
| Case 12 | 0.735 | 0.683 | 3.16 | 2.40 | 2.03 | 0.818 | 0.789 | 68 | M | ADD | + | BF2 |
| Case 13 | 0.753 | 0.768 | 1.15 | 1.98 | 1.79 | 0.641 | 0.657 | 74 | F | bvFTD | + | BF2 |
| Case 14 | 0.842 | 0.817 | 3.25 | 2.21 | 1.99 | 0.921 | 0.926 | 65 | M | ADD | + | BF2 |
| Case 15 | 0.778 | 0.776 | 1.83 | 1.19 | 1.18 | 0.850 | 0.836 | 71 | F | MCI | + | BF2 |
| Case 16 | 0.779 | 0.785 | 1.17 | 1.14 | 1.19 | 0.154 | 0.126 | 77 | F | CN | - | PREVENT-AD |
| Case 17 | 0.778 | 0.785 | 1.41 | 1.06 | 1.08 | 0.162 | 0.301 | 61 | F | ADD | + | UCSF |
| Case 18 | 0.774 | 0.772 | 1.24 | 1.14 | 1.22 | 0.190 | -0.0450 | 68 | F | CN | - | PREVENT-AD |
| Case 19 | 0.715 | 0.679 | 2.55 | 1.54 | 1.21 | 0.205 | 0.0409 | 73 | F | ADD | + | BF2 |
| Case 20 | 0.695 | 0.698 | 1.38 | 1.14 | 1.16 | 0.175 | 0.261 | 65 | M | CN | - | PREVENT-AD |
| High-performing example cases (Supplementary Fig. 5) | | | | | | | | | | | | |
| Case 21 | 0.889 | 0.889 | 1.21 | 1.21 | 1.23 | 0.763 | 0.745 | 75 | F | CN | + | A4 |
| Case 22 | 0.794 | 0.794 | 1.23 | 1.23 | 1.23 | 0.650 | 0.658 | 72 | F | CN | - | PREVENT-AD |
| Case 23 | 0.834 | 0.832 | 1.86 | 1.86 | 1.59 | 0.829 | 0.841 | 81 | M | ADD | + | BF2 |
| Case 24 | 0.838 | 0.835 | 1.32 | 1.33 | 1.43 | 0.941 | 0.939 | 63 | M | ADD | + | BF2 |
| Case 25 | 0.829 | 0.801 | 2.05 | 2.07 | 1.68 | 0.815 | 0.764 | 71 | F | LPA | + | UCSF |
| Case 26 | 0.837 | 0.831 | 2.21 | 2.13 | 1.90 | 0.920 | 0.902 | 78 | M | ADD | + | BF2 |
| Case 27 | 0.806 | 0.798 | 1.48 | 1.39 | 1.30 | 0.927 | 0.881 | 73 | M | ADD | + | BF2 |
| Case 28 | 0.847 | 0.844 | 1.99 | 1.90 | 1.52 | 0.918 | 0.911 | 85 | M | ADD | + | BF2 |
| Case 29 | 0.759 | 0.727 | 1.70 | 1.59 | 1.37 | 0.915 | 0.906 | 54 | M | MCI | + | BF2 |
| Case 30 | 0.811 | 0.807 | 1.40 | 1.61 | 1.46 | 0.934 | 0.942 | 79 | M | ADD | + | UCSF |
| Representative example cases from cohorts without plasma p-tau217 (Supplementary Fig. 7) | | | | | | | | | | | | |
| Case 31 | - | 0.859 | 1.22 | - | 1.24 | - | 0.789 | 70 | M | CN | + | A04 |
| Case 32 | - | 0.873 | 1.52 | - | 1.30 | - | 0.897 | 77 | F | ADD | + | A05 |
| Case 33 | - | 0.803 | 1.21 | - | 1.24 | - | 0.611 | 78 | F | SCD | + | A08 |
| Case 34 | - | 0.864 | 2.16 | - | 2.09 | - | 0.889 | 62 | F | ADD | + | LLCF |
| Case 35 | - | 0.827 | 2.21 | - | 1.89 | - | 0.874 | 58 | F | ADD | + | LZAX |
| Case 36 | - | 0.849 | 1.20 | - | 1.16 | - | 0.768 | 77 | M | CN | N/A | BACS |
| Case 37 | - | 0.832 | 1.41 | - | 1.46 | - | 0.852 | 84 | F | CN | + | OASIS |

**Supplementary Table 6: Performance metrics and basic demographic characteristics for example cases.**

**Supplementary Table 7: Demographics of BF1 individuals without tau-PET.**

|  | **BF1 individuals without tau-PET (progression to dementia analysis)** |
| --- | --- |
| **Number of participants** | 358 |
| **Age, mean (std) years** | 72.7 (5.55) |
| **Sex n (%) female** | 220 (61.5) |
| ***APOE* ε4 alleles*, n (%)** |  |
| 0 | 240 (67.2) |
| 1 | 100 (28.0) |
| 2 | 17 (4.76) |
| **Cognitive status, n (%)** |  |
| NC | 214 (59.8) |
| SCD | 144 (40.2) |
| **Aβ positive*, n (%)** | 158 (44.1) |
| **Plasma p-tau217*, mean (std) pg/ml** | 0.209 (0.119) |
| **Follow-up duration, mean (std) months** | 78.2 (29.5) |
| **Progression to all-cause dementia, n (%)** | 50 (16.2) |

*missing data for some participants

| Example Case | True Braak I-IV load | Synthetic Braak I-IV load (p-tau217) | Synthetic Braak I-IV load (CSF MTBR-tau243) | Synthetic Braak I-IV load (p-tau217 & CSF MTBR-tau243) | Spatial correlation (p-tau217) | Spatial correlation (CSF MTBR-tau243) | Spatial correlation (p-tau217 & CSF MTBR-tau243) | Age | Sex | Diagnosis | Aβ status | Cohort |
| --- | --- | --- | --- | --- | --- | --- | --- | --- | --- | --- | --- | --- |
| Representative example cases that improved with a combined MRI, age, plasma p-tau217 and CSF MTBR-tau243 model (Fig. 7) | | | | | | | | | | | | |
| Case 38 | 1.61 | 1.17 | 1.45 | 1.55 | 0.69 | 0.82 | 0.84 | 68 | M | ADD | + | BF2 |
| Case 39 | 2.17 | 1.90 | 1.69 | 2.02 | 0.77 | 0.77 | 0.70 | 81 | M | MCI | + | BF2 |
| Case 40 | 1.16 | 1.54 | 1.52 | 1.23 | 0.49 | 0.49 | 0.75 | 75 | F | svPPA | - | BF2 |

**Supplementary Table 8: Performance metrics and basic demographic characteristics for example cases with CSF MTBR-tau243 as input.**

**Supplementary Table 9: Model performance comparing plasma p-tau217, plasma p-tau181 and CSF MTBR-tau243 in single and combined models.** Analysis made on a subset of the test sample with all variable available (n=178).

| **Model input** | **Mean (std) SSIM** | **MAE (std) Braak I-IV load** | **Mean (std) within-subject spatial correlation R** | **R Braak I-IV load** | **P-value FDR  R Braak I-IV (compared against)** |
| --- | --- | --- | --- | --- | --- |
| Single biomarker models | | | | | |
| MRI, age, plasma p-tau217 | 0.800 (0.041) | 0.140 | 0.795 (0.132) | 0.848 | - |
| MRI, age, plasma p-tau181 | 0.759 (0.042) | 0.145 | 0.798 (0.123) | 0.840 | 0.712  (MRI, age, plasma p-tau217) |
| MRI, age, CSF MTBR-tau243 | 0.800 (0.042) | 0.140 | 0.804 (0.121) | 0.857 | 0.712  (MRI, age, plasma p-tau217) |
| Combined biomarker models | | | | | |
| MRI, age, plasma p-tau217, plasma p-tau181 | 0.787 (0.044) | 0.145 | 0.798 (0.119) | 0.839 | - |
| MRI, age, plasma p-tau217, CSF MTBR-tau243 | 0.752 (0.049) | 0.124 | 0.830 (0.0946) | 0.890 | **0.018** (MRI, age, plasma p-tau217, plasma p-tau181) |

**Supplementary Table 10: Definitions of imaging composites from FreeSurfer parcellation and segmentation.**

| **Composite name** | **Included regions** |
| --- | --- |
| Braak I-II | Entorhinal. |
| Braak I-IV | Entorhinal; parahippocampal; fusiform; inferiortemporal; middletemporal; amygdala. |
| Braak V-VI | Caudalanteriorcingulate; cuneus; inferiorparietal; isthmuscingulate; lateraloccipital; lateralorbitofrontal; lingual; medialorbitofrontal; paracentral; parsopercularis; parsorbitalis; parstriangularis; pericalcarine; postcentral; posteriorcingulate; precentral; precuneus; rostralanterioscingulate; rostralmiddlefrontal; superiorfrontal; superiorparietal; superiortemporal; supramarginal; frontalpole ;temporalpole; transversetemporal; insula. |
| MRI AD signature | Entorhinal; fusiform; inferiortemporal; middletemporal. |
| T_MTL_ | Entorhinal; amygdala. |
| T_NEO_ | Middletemporal; inferiortemporal; |
