## Supplementary Figures for "Generating synthetic tau-PET scans in Alzheimer’s disease from MRI, blood biomarkers and demographics with deep learning"

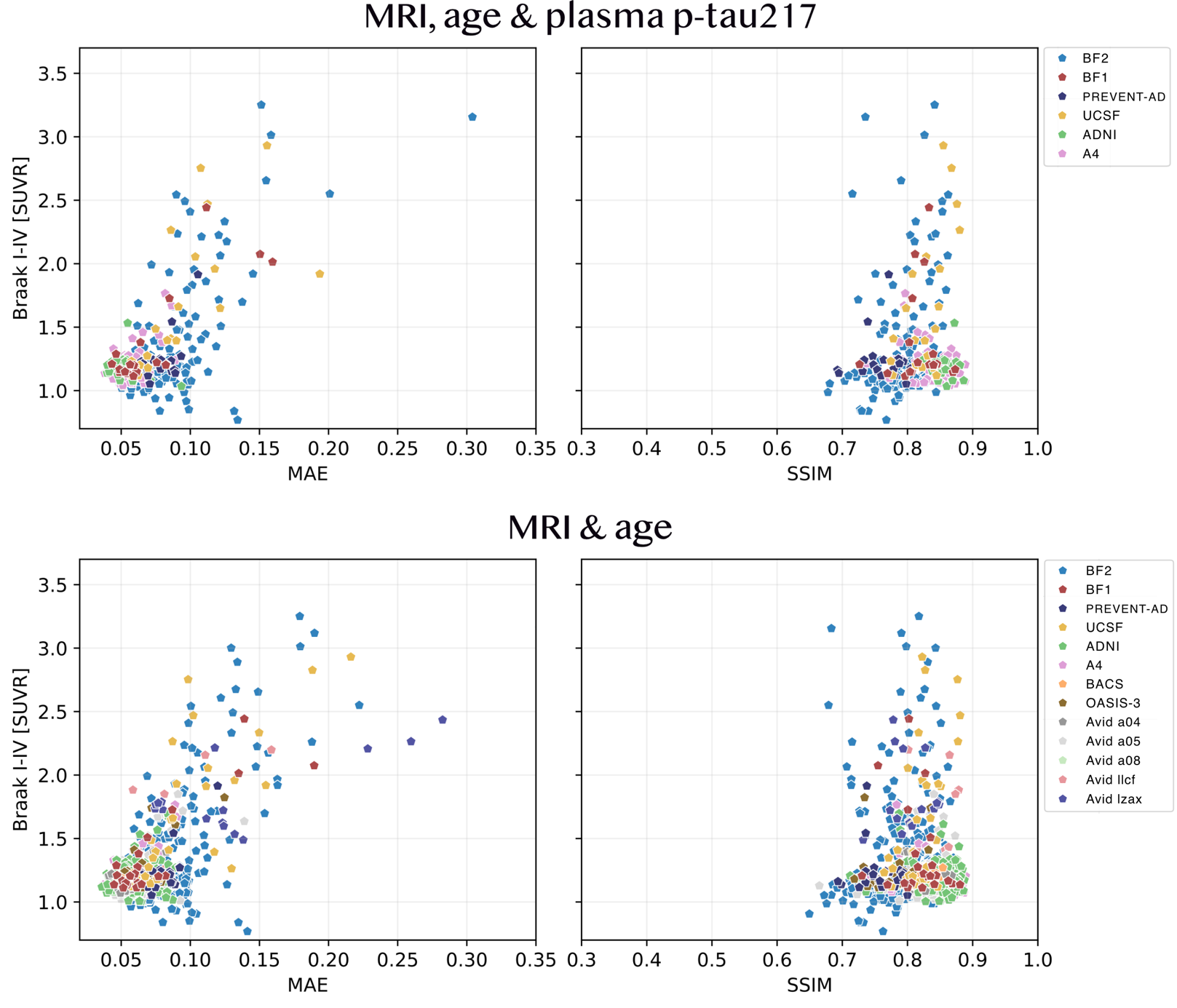

**Supplementary Figure 1: Image quality assessment in the test set.** Scatter plots of MAE and SSIM versus true Braak I-IV load.

Abbreviations: Structural similarity index measure (SSIM), mean absolute error (MAE).

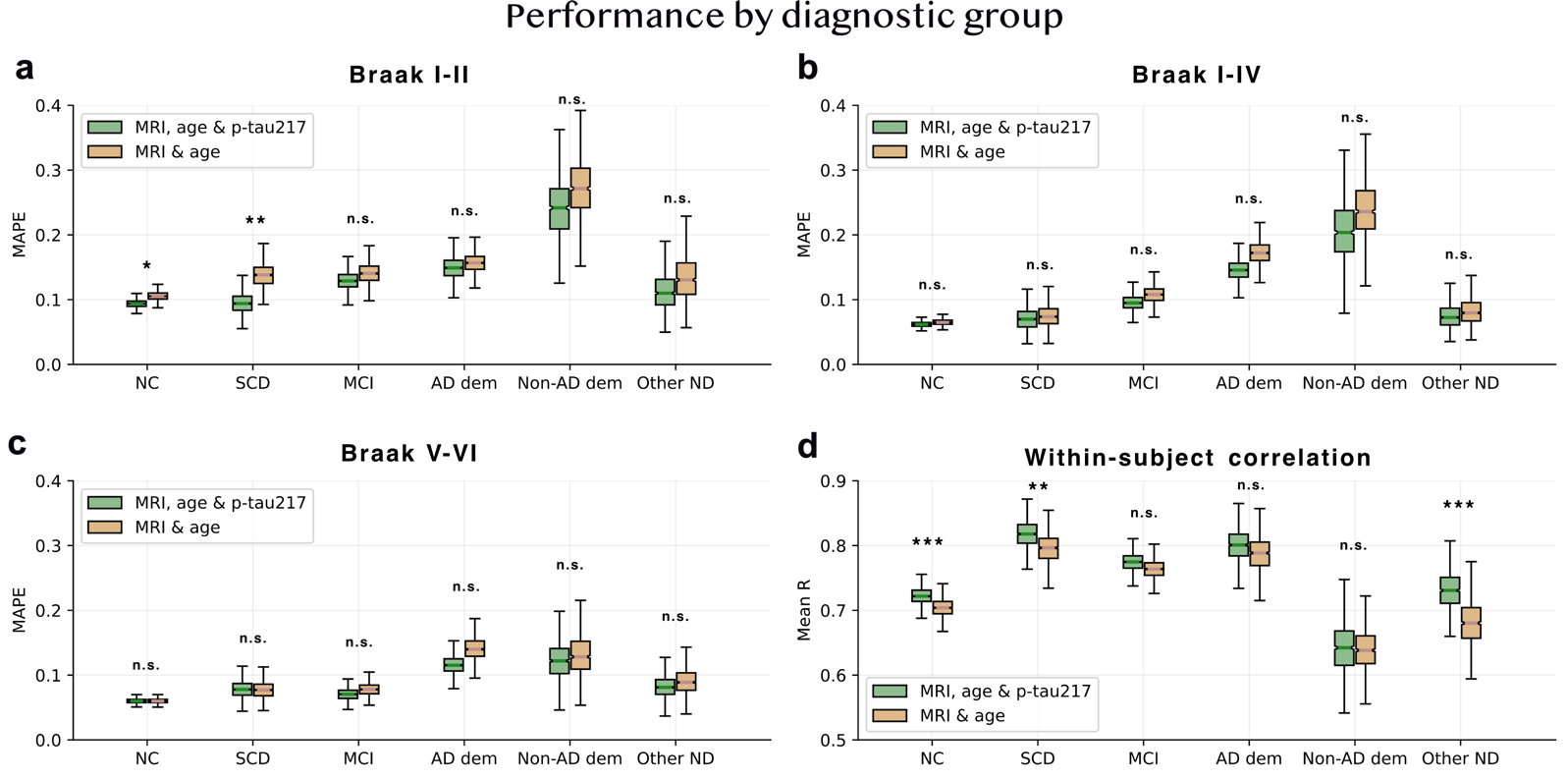

**Supplementary Figure 2: Tau load and spatial distribution performance across diagnostic groups.**

Non-AD dementia includes Frontotemporal dementia, Lewy body dementia, Vascular dementia, and Posterior cortical atrophy. Other neurological disorder includes Parkinson's disease, Progressive supranuclear palsy, Corticobasal syndrome, multiple system atrophy, traumatic brain injury, chronic traumatic encephalopathy, and bipolar disorder. Number of participants in each group: n_CN_ = 156, n_SCD_ = 22, n_MCI_ = 49, n_ADD_ = 46, n_non-ADD_ = 18, n_OND_ = 27.

Abbreviations: mean absolute percentage error (MAPE), normal cognition (NC), subjective cognitive decline (SCD), mild cognitive impairment (MCI), Alzheimer’s disease (AD), neurological disorder (ND).

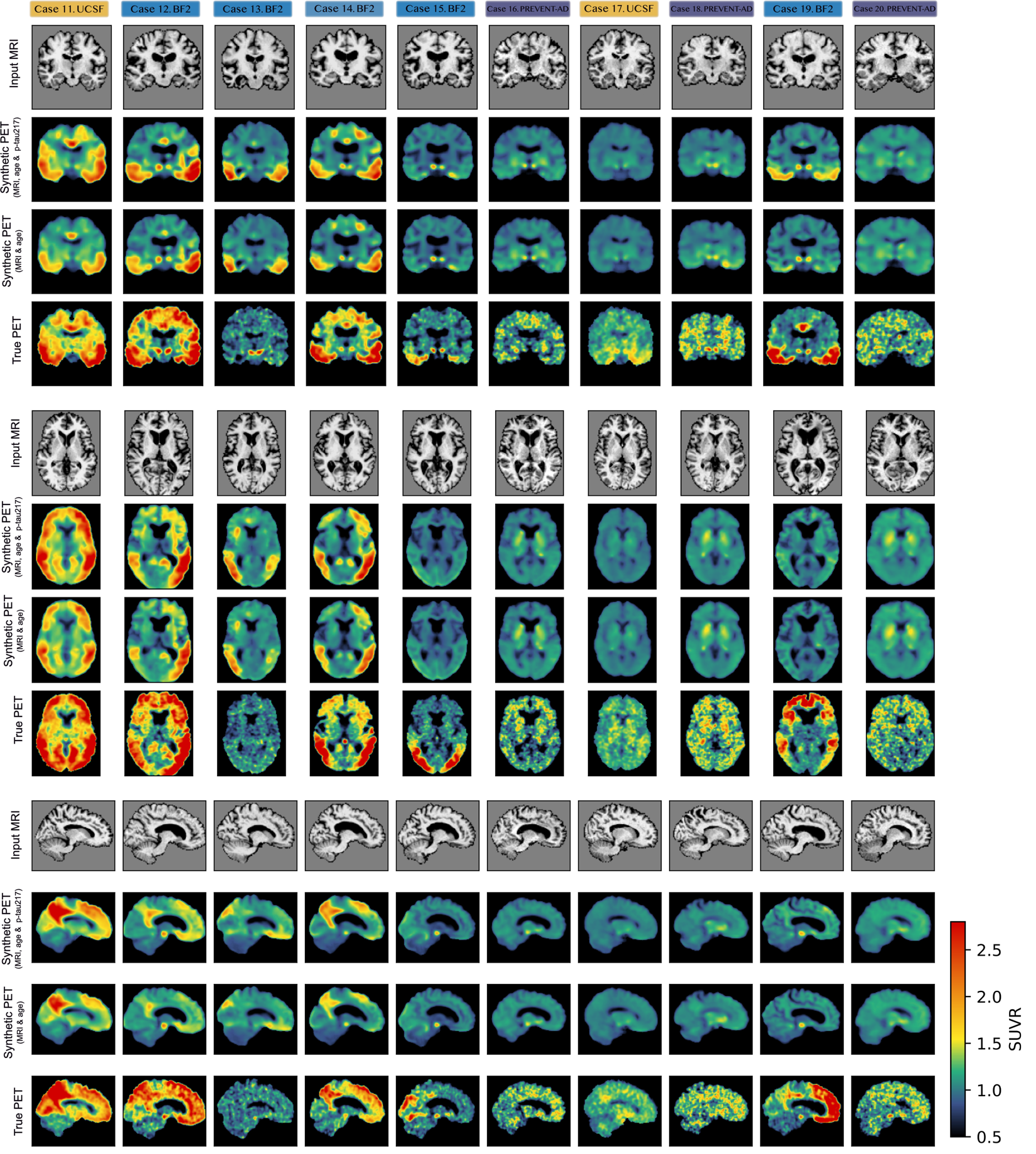

**Supplementary Figure 3: Ten low-performing example cases.** The input MRI (first row), synthetic tau-PET based on MRI, age and plasma p-tau217 (second row), synthetic tau-PET based on MRI and age (third row), and true tau-PET (fourth row). Corresponding performance metrics and basic demographic information for case are presented Supplementary Tab. 6.

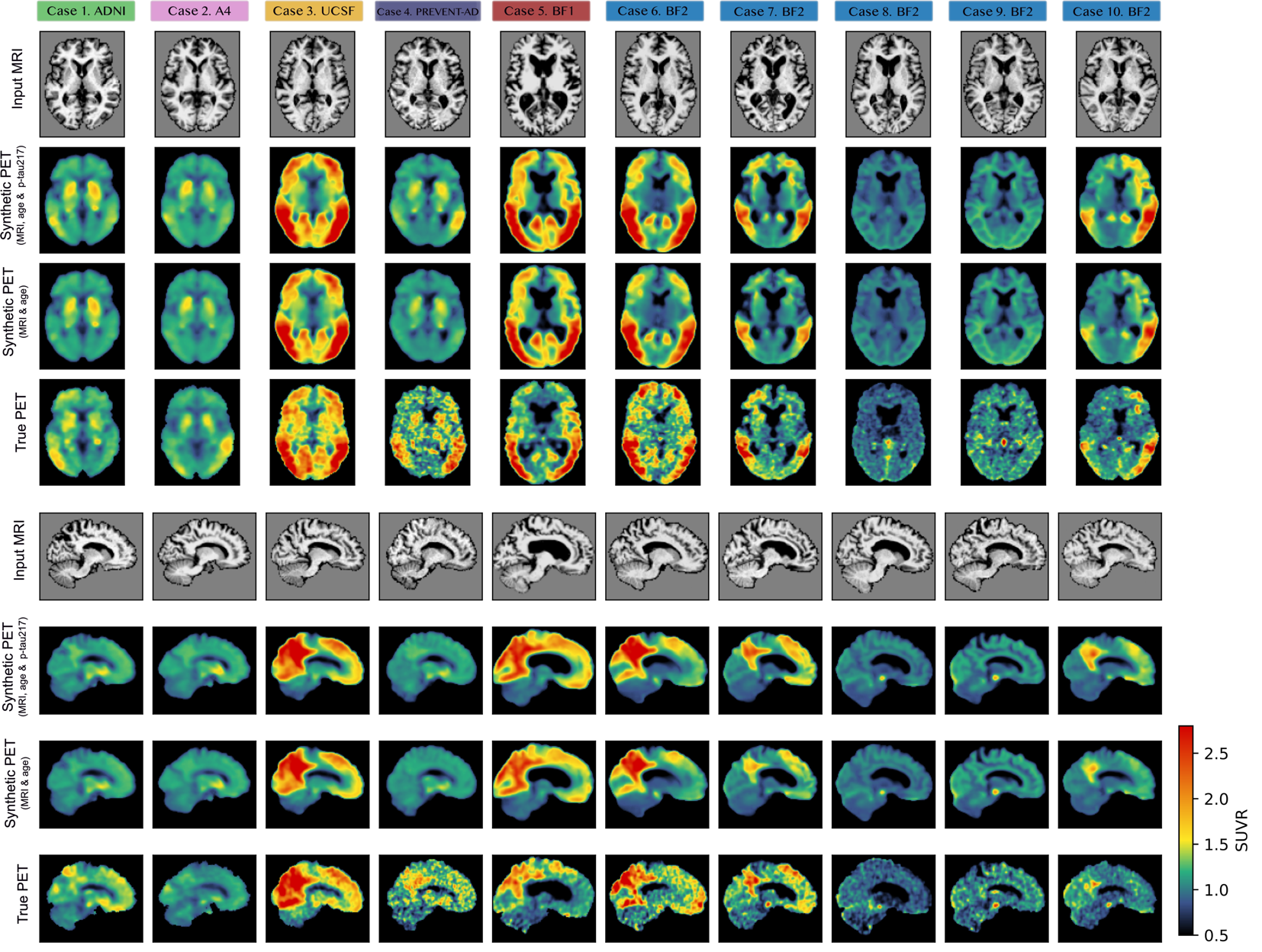

**Supplementary Figure 4: Ten representative example cases from axial and sagittal views (same as Figure 5).** The input MRI (first row), synthetic tau-PET based on MRI, age and plasma p-tau217 (second row), synthetic tau-PET based on MRI and age (third row), and true tau-PET (fourth row). Corresponding performance metrics and basic demographic information for case are presented Supplementary Tab. 6.

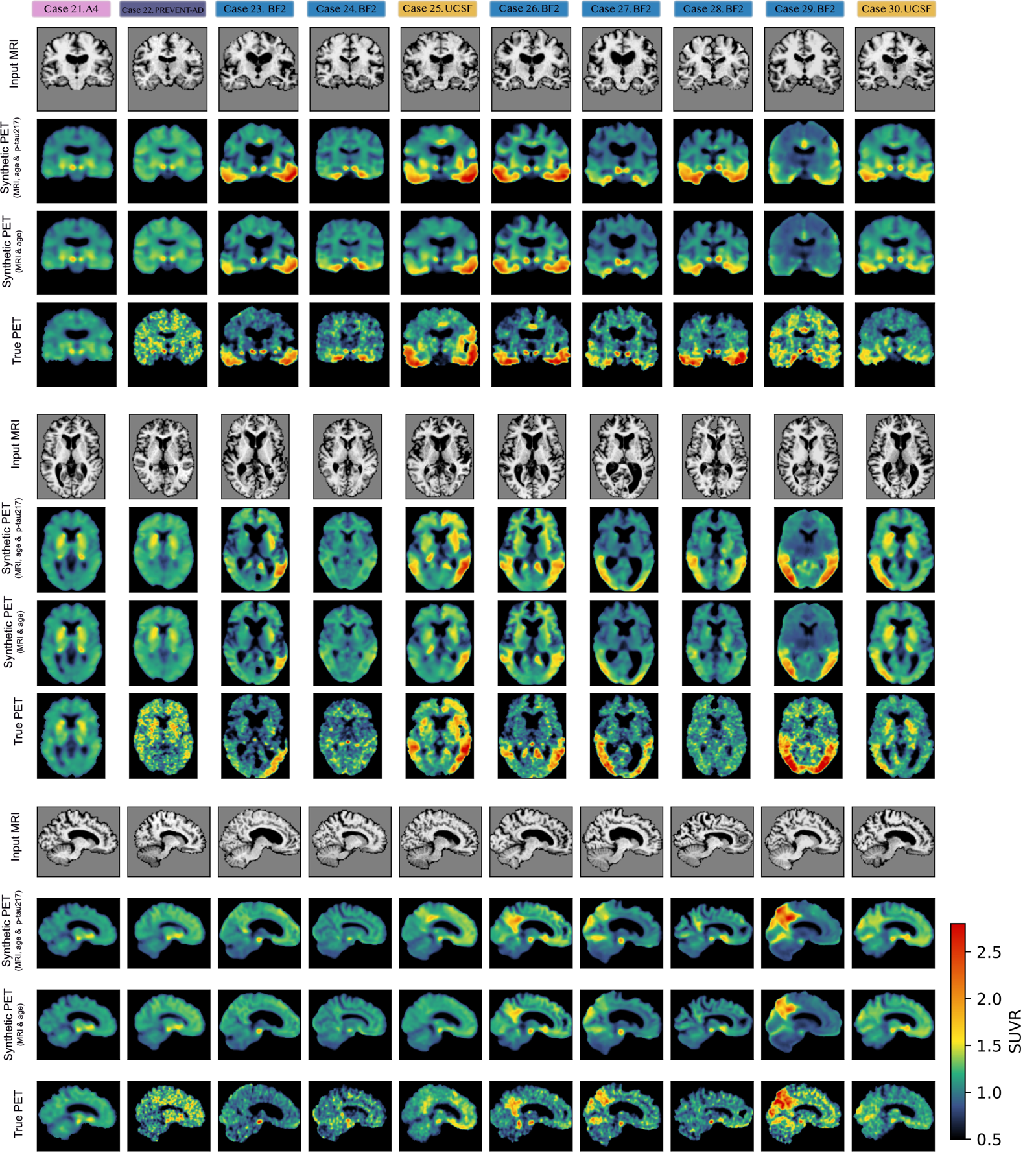

**Supplementary Figure 5: Ten high-performing example cases.** The input MRI (first row), synthetic tau-PET based on MRI, age and plasma p-tau217 (second row), synthetic tau-PET based on MRI and age (third row), and true tau-PET (fourth row). Corresponding performance metrics and basic demographic information for case are presented Supplementary Tab. 6.

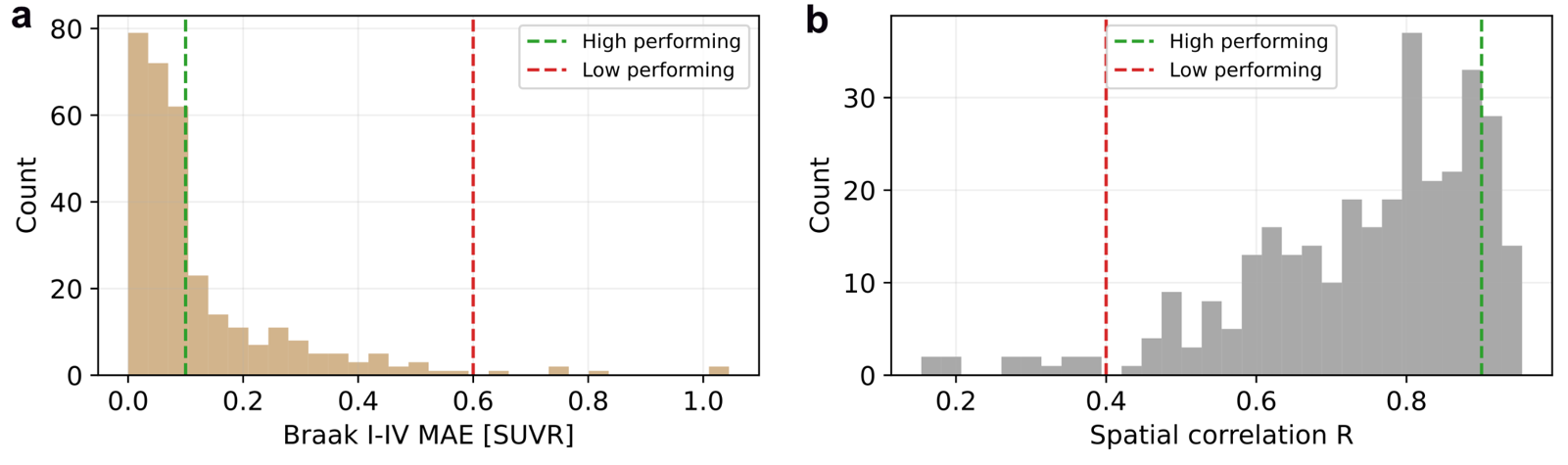

**Supplementary Figure 6: Test set distribution of performance metrics used to select high/low-performing cases.** a) Braak I-IV MAE, with high-performing cases showing errors smaller than 0.1 SUVR (green line) and low-performing cases higher than 0.6 SUVR (red line). and b) spatial correlation, with high-performing cases showing correlation higher than R=0.9 (green line) and low-performing cases lower than R=0.4 SUVR (red line).

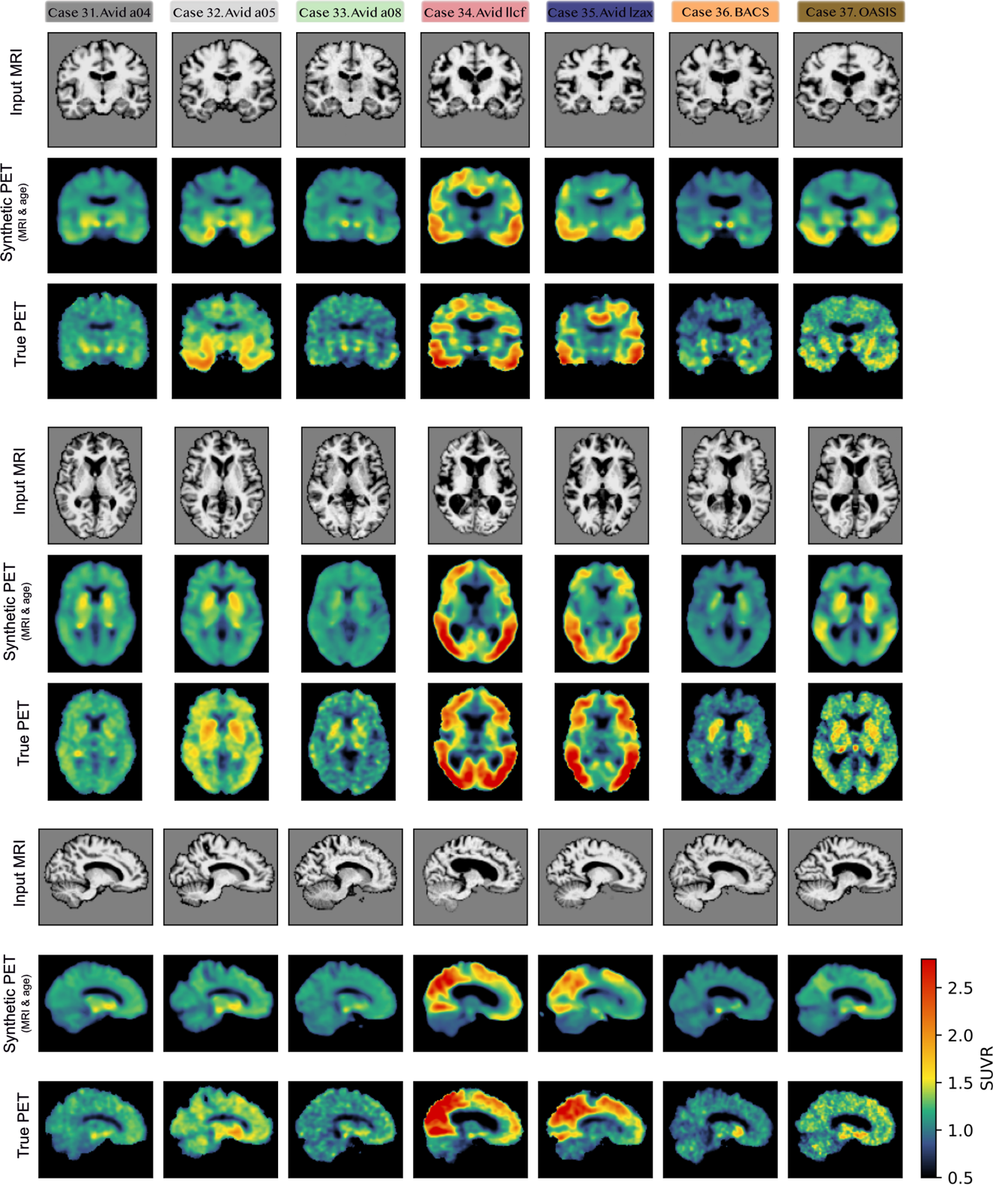

**Supplementary Figure 7: Example cases from cohorts without plasma p-tau217 available.** The input MRI (first row), synthetic tau-PET based on MRI and age (second row), and true tau-PET (third row). Corresponding performance metrics and basic demographic information for case are presented Supplementary Tab. 6.
