## Supplementary Note 1 for "Generating synthetic tau-PET scans in Alzheimer’s disease from MRI, blood biomarkers and demographics with deep learning"

**Voxel-wise synthetic tau-PET errors differ by diagnosis and tracer**

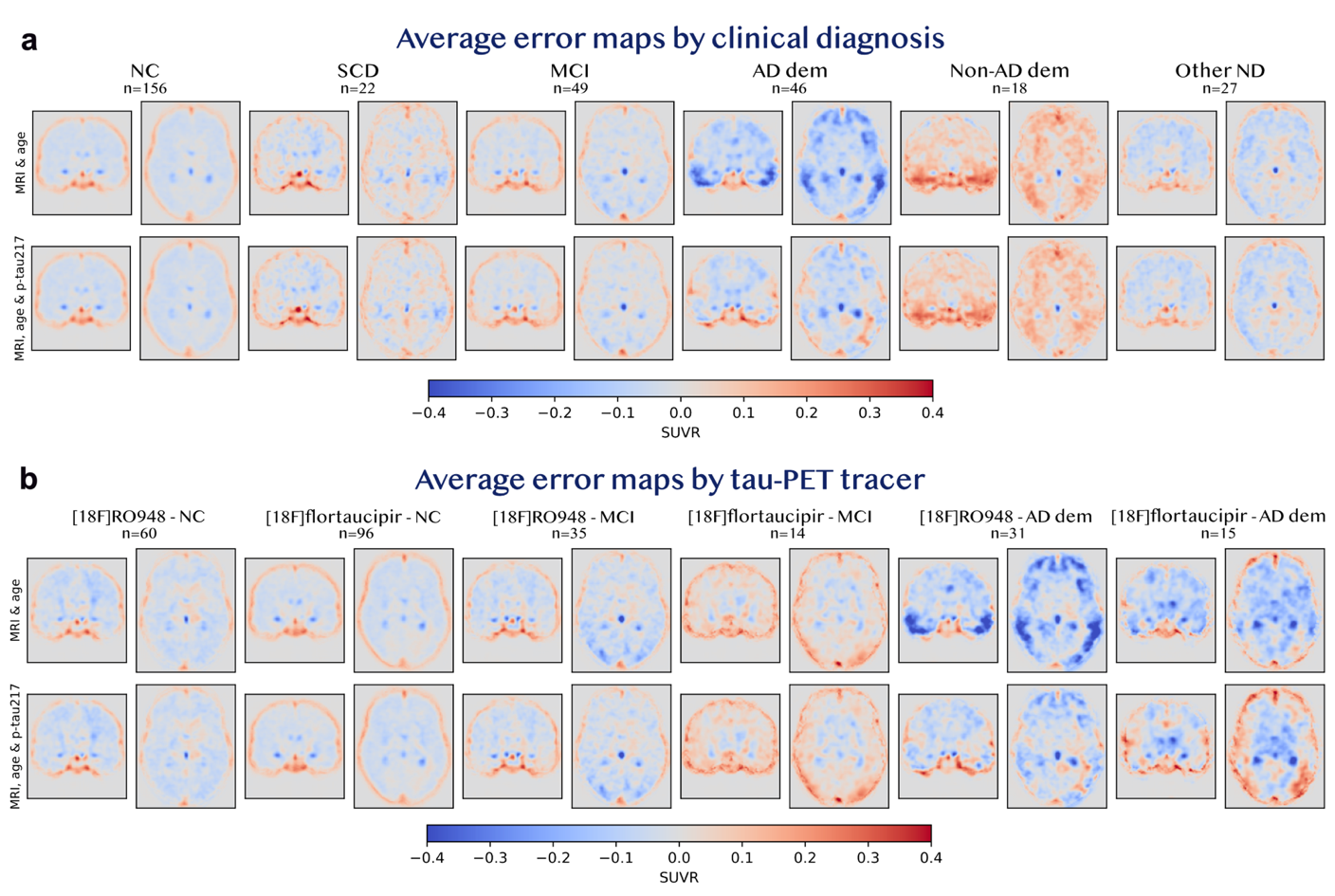
To visualize systematic error biases, we averaged voxel-wise errors between synthetic and true tau-PET across subjects, stratified by clinical diagnosis and tau-PET tracer (Fig. SN1). Synthetic scans consistently underestimated diffuse neocortical uptake, as well as signal in regions known for off-target binding, particularly the choroid plexus and basal ganglia (previously reported higher for [^18^F]flortaucipir than [^18^F]RO948).^1,2^ Another recurring pattern was a systematic overestimation bias along the cortical edge, likely due to off-target binding in the skull/meninges (previously reported higher for [^18^F]RO948 than [^18^F]flortaucipir^3^) and the smoother brain boundary in the synthetic scans compared to true (apparent in Fig. 5; could be mitigated by additional post-processing). Comparing clinical diagnoses, average errors were largest for the AD dementia and non-AD dementia groups. The inclusion of plasma p-tau217 appeared to partially corrected underestimation of tau in temporal, parietal and frontal regions in participants with AD dementia, but had less effect on other diagnostic groups. Plasma p-tau217 generally increased signal in temporal and parietal regions for the AD-dementia group, an adjustment that alleviated underestimation for [^18^F]RO948 images but introduced mild overestimation for [^18^F]flortaucipir images.

**Figure SN1: Systematic error biases stratified by clinical diagnosis and tau-PET tracer.** Voxel-wise error in the test set averaged across a) diagnostic groups and b) diagnostic groups and tau-PET tracers. Overestimations of signal in synthetic PET compared to true PET colored by red, and underestimations by blue. The SCD, non-AD dementia and other ND groups were not stratified by tracer due to small sample sizes (n ≤ 5 for [^18^F]flortaucipir). Abbreviations: Standardized uptake value ratio (SUVR), normal cognition (NC), mild cognitive impairment (MCI), Alzheimer’s disease (AD), neurological disorder (ND), dementia (dem)
